# Population-scale analysis reveals limited and non-generalizable associations between the gut microbiome and obesity in Asian adults

**DOI:** 10.64898/2026.08.12.26358109

**Authors:** Jonathan J.Y. Teo, Benjamin C. H. Lam, Shaun Hong Chuen How, Ruwen Zhou, Sunny H. Wong, John C. Chambers, Niranjan Nagarajan

**Author notes:** Co-first Authors (Jonathan J.Y. Teo, Benjamin C. H. Lam).

## Abstract

**Background:** The gut microbiome has been widely studied in the context of obesity, and yet the reported associations vary widely across populations and analytical approaches. In Asian populations where the prevalence of obesity is rapidly rising, the extent to which gut microbiome features could associate with adiposity in a robust and generalizable manner remains unclear.

**Methods:** Population-scale shotgun metagenomic data was generated for adults (n=871) from the Health for Life in Singapore (HELIOS) cohort, comprising ethnic Chinese, Malay, and Indian participants. Integrated taxonomic, functional, and machine-learning–based analyses were used to assess associations between gut microbiome features and obesity, adjusting for demographic covariates and evaluating for robustness across multiple statistical frameworks.

**Results:** Global microbiome structure exhibited weak separation by body mass index (BMI), with enterotype-like clustering providing limited discriminatory power for obesity status. Differential abundance analyses identified a small number of method-dependent taxa and pathways, with only limited recurrence across methods. Supervised machine learning models trained on taxonomic profiles achieved modest predictive performance, particularly for intermediate BMI classes, and did not reveal robust microbial signatures beyond those detected by univariate analyses.

**Conclusions:** Our study highlights the importance of large-scale, multi-framework analyses for distinguishing robust microbiome–phenotype associations from weak, method-dependent signals. Together, our findings emphasize that obesity-associated microbiome signatures may be too weak, diffuse, and insufficient to explain adiposity in Asian populations.

## Introduction

Over the past several decades, the global prevalence of being overweight and obese has increased substantially, affecting hundreds of millions of individuals and contributing to a major rise in non-communicable diseases (NCDs) worldwide^1,2^. In Asia, rapid urbanization and economic development have coincided with major shifts in dietary patterns, physical activity, and overall lifestyle, fueling a regional nutrition transition and rising obesity prevalence across both high-income and low- and middle-income settings^3–5^. Because obesity is a major risk factor for type 2 diabetes, cardiovascular disease, and several cancers^3,4^, understanding its biological and environmental contributors has become an urgent research priority, particularly in urban, multi-ethnic populations where lifestyle exposures increasingly converge while health outcomes remain heterogeneous^4,5^.

The gut microbiome has emerged as a candidate contributor to obesity, following repeated reports that adiposity is associated with differences in microbial community structure, ecological richness, and metabolic potential across multiple sequencing strategies. However, despite extensive study, the specific microbial features linked to obesity have varied substantially across cohorts, sequencing strategies, and analytical frameworks. Early 16S rRNA gene studies reported compositional differences between individuals with obesity and lean controls, including shifts in taxa such as *Bacteroides, Prevotella, Ruminococcus,* and *Bifidobacterium*, and in some cohorts, reduced alpha diversity or overall community richness^6,7^. These early observations helped establish the-idea that obesity may be accompanied not only by changes in individual taxa but also by broader perturbations in gut ecosystem organization, though cohort sizes often limited the ability to fully account for confounders. Subsequent metagenomic studies extended this framework by emphasizing microbial gene content and functional capacity, including pathways related to carbohydrate utilization, fermentation, short-chain fatty acid production, branched-chain amino acid biosynthesis, bile acid transformation, oxidative stress responses, and other energy-harvesting or host-interactive functions^8–10^. In parallel, the field increasingly recognized that some early taxonomic signatures, particularly the *Firmicutes:Bacteroidetes* ratio, were influential but not consistently reproducible across cohorts, highlighting the context dependence and sensitivity to confounders in identifying obesity-associated gut microbial patterns^6,8,11^. Subsequent meta-analyses across multiple cohorts have shown that effect sizes for individual obesity-associated taxa are typically small and not robustly reproducible^12–14^, suggesting that prior signals may be shaped by cohort heterogeneity, methodology, or context. Evidence for a causal contribution of the microbiome has come primarily from gnotobiotic mouse studies. Colonization of germ-free mice with conventional microbiota increased host adiposity and energy harvest relative to germ-free controls^15^, while transplantation of microbiota from obese donors, including obesity-discordant human twins, promoted greater fat gain and metabolic impairment in recipient animals under controlled dietary conditions^16^. Together, these findings support the growing evidence that the gut microbiome can modulate host energy balance and may contribute to obesity.

Critically, whether the microbiome contributes meaningfully to obesity in human populations depends not only on whether associations are statistically detectable, but on the magnitude of microbiome effects relative to known host and environmental determinants of adiposity, and after accounting for diverse technical and biological confounders^17^. The strongest evidence for causality has come from gnotobiotic mouse studies, which support microbiome-mediated effects on host adiposity but do not necessarily translate directly to human populations^16^. A limited number of interventional studies, including using biotherapeutic approaches, support the relevance of specific microbes or microbial functions, but such effects may not translate into strong or reproducible population-scale associations^18–20^. Moreover, much of the human evidence base has been shaped largely by Western and other select cohorts, and it remains unclear whether the microbial features most often linked to obesity are reproducible in multi-ethnic Asian populations, where dietary exposures, adiposity distribution, and metabolic risk profiles differ substantially. More broadly, population-based studies have shown that gut microbiome composition varies across Western and Asian populations, even after accounting for shared geography in some settings, supporting the idea that ethnicity, diet, urbanization, and other contextual exposures can shape baseline microbial structure and may influence the generalizability of obesity-associated signals^1^. Existing studies in Chinese populations, including Han Chinese intervention cohorts and pediatric obesity studies, provide valuable early signals but have generally been limited in size or study design^23,24^. At the analytical level, prior work has also relied heavily on differential-abundance testing within a single statistical framework, which may be sensitive to model choice and less able to detect multivariate or non-linear structure in microbiome data^25^. By contrast, relatively few obesity-related microbiome studies have combined differential-abundance testing with machine-learning approaches to evaluate whether microbial features are robust enough for prediction or stratification^26,27^. Robust inference therefore requires comparing results across multiple analytical paradigms, including univariate and multivariate, linear and non-linear approaches, which has rarely been done systematically in obesity-microbiome research.

In this study, we re-examined gut microbiome-obesity associations in a large, multi-ethnic Asian cohort (ethnic Chinese, Malay, and Indian) residing in Singapore, where the population share a highly urbanized environment, food supply, and healthcare system, which reduces some sources of environmental heterogeneity that can complicate cross-country comparisons. We first characterized population-level microbiome structure and enterotype-like clustering to assess their associations with BMI and obesity. We then evaluated obesity-associated microbial signals using complementary differential abundance and machine learning approaches, followed by pathway-level analyses of microbial function while adjusting for demographic covariates. Across these analyses, we observed generally weak associations between the gut microbiome and obesity, characterized by modest effect sizes, limited concordance across analytical methods, and substantial overlap between BMI groups. Together, our findings indicate that obesity-related microbiome signatures explain only a limited fraction of adiposity-associated variation at the population scale.

## Results

### Microbiome structure and enterotype-like clustering show limited associations with obesity

To evaluate whether gut microbiome features are robustly associated with obesity in an Asian population, we generated and analyzed shotgun metagenomic profiles from 871 adults enrolled in the Health for Life in Singapore (HELIOS) cohort together with standardized demographic and anthropometric measurements. This cohort provided a large, multi-ethnic Southeast Asian setting in which obesity-related microbiome variation could be assessed across ethnic Chinese, Malay, and Indian participants living within the shared urban environment of Singapore. The cohort was 38% male and 62% female, and was predominantly ethnic Chinese (82.9%), with smaller proportions of ethnic Indian (10.7%) and ethnic Malay (6.4%) participants. Participants ranged in age from 30.3 to 77.0 years, with a mean age of 49.9±11.1 years. The mean BMI was 24.0±4.2 kg/m², and mean waist circumference was 82.0±11.2 cm. Using WHO Asian BMI cutoffs, 47.2% were normal (<22.9 kg/m²), 36.4% were pre-obese (23.0–27.4 kg/m²), and 16.4% were obese (≥27.5 kg/m²; **Methods**). This distribution captures substantial variation in adiposity within a shared urban, multi-ethnic setting.

Stool samples were collected from participants for deep shotgun metagenomic sequencing, generating >19 billion reads and >20 million reads on average per sample (n=871 libraries; **Methods**). All samples were consistently processed to generate taxonomic profiles, where unsupervised ordination analysis at the species-level revealed substantial inter-individual variability but no strong global separation by BMI or ethnicity (**Fig. 1A**; **Supplementary Fig. 1**). This pattern was also robust across alternative distance metrics and ordination settings (**Supplementary Fig. 1**), with samples from different BMI categories and ethnic groups remaining broadly intermingled. These results suggest that population-wide gut microbiome structure is only weakly associated, at most, with adiposity and ethnicity in this cohort.

**Figure 1.**
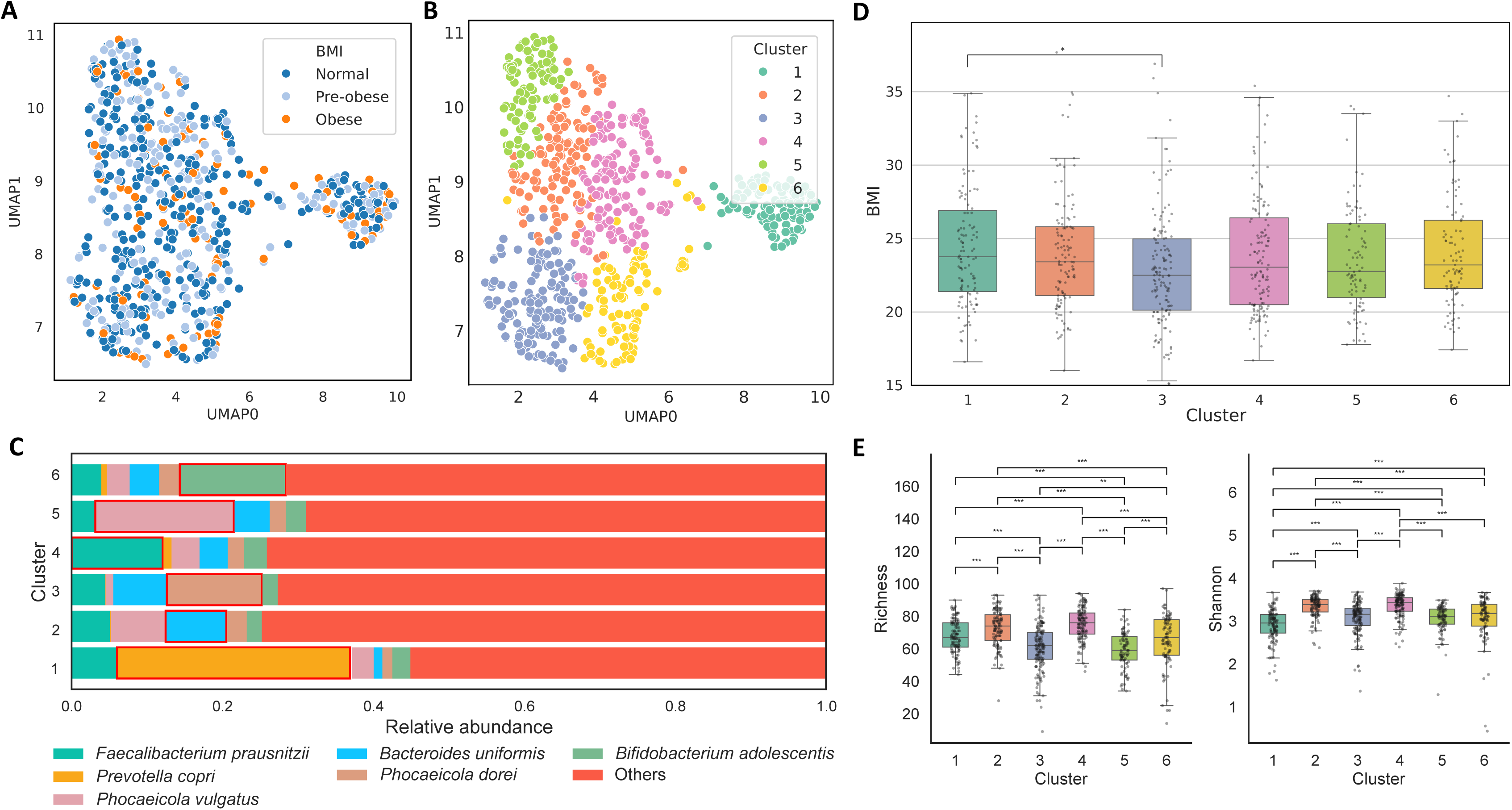
Global gut microbiome structure shows reproducible community patterns but weak association with obesity. **(A)** Uniform Manifold Approximation and Projection (UMAP) of species-level gut microbiome profiles using Bray–Curtis distance, colored by Asian-specific BMI category (normal, pre-obese, obese). No clear separation by BMI category was observed. Permutational multivariate analysis of variance (PERMANOVA; 9,999 permutations) indicated that BMI explained only a very small fraction of total variation (R²=0.0069, P=0.0001), with similarly modest contributions from ethnicity (R²=0.0079), age (R²=0.0045), and sex (R²=0.0080) (**Supplementary Table 1**). **(B)** The same UMAP embedding colored by cluster labels from unsupervised spectral clustering (k=6), selected based on elbow and silhouette criteria (**Methods**; **Supplementary** Fig. 2). Clusters reflect gradients in dominant commensal taxa rather than discrete phenotype-specific groupings. **(C)** Mean relative abundance of six prevalent gut taxa across spectral clusters, with remaining taxa aggregated as “Others.” Cluster differentiation is driven by shifts in the relative dominance of common commensals, including *Faecalibacterium prausnitzii, Prevotella copri, Phocaeicola vulgatus, Bacteroides uniformis, Phocaeicola dorei,* and *Bifidobacterium adolescentis*, rather than by obesity-specific taxa. **(D)** Distribution of BMI across clusters. Although median BMI differs modestly between some clusters, the distributions overlap extensively. Pairwise comparisons were performed using two-sided Mann-Whitney U tests with Benjamini-Hochberg correction, with adjusted P<0.05 indicated. **(E)** Alpha diversity (species richness and Shannon diversity) varies significantly across spectral clusters (pairwise Mann-Whitney U tests with Benjamini-Hochberg correction; *** q < 0.001, ** q < 0.01). Clusters 2 and 4 show consistently higher diversity than clusters 3 and 5, reflecting compositional gradients that track dominant-taxon shifts rather than BMI category.

Consistent with the ordination patterns, permutational multivariate analysis of variance (PERMANOVA) showed that obesity-related variation explained only a very small fraction of overall gut microbiome composition. BMI category accounted for less than 1% of the total variance (R²=0.0069, P=0.0001), indicating that global community structure was weakly associated with adiposity (**Fig. 1A; Supplementary Table 1**). Other host factors, including ethnicity (R²=0.0079, P=0.0001), age (R²=0.0045, P=0.0004), and sex (R²=0.0080, P=0.0001), explained similarly modest proportions of variance. This near-null result was not specific to BMI. Repeating the analysis with central-adiposity measures, in a cohort matched to the BMI analysis and adjusted for age, sex, and ethnicity, left the overall interpretation unchanged: waist circumference explained essentially none of the species-level compositional variance (R²=0.0043, P=0.0004), as did waist-to-hip ratio (WHR, R²≈0.0027, P=0.0238) and waist-to-height ratio (WHtR, R²≈0.0040, P=0.0015). Each matched the equally negligible continuous BMI estimate in the same matched cohort (R²=0.0056; **Supplementary Table 1**), indicating the absence of a global compositional signal that holds across adiposity metrics, rather than being an artefact of BMI-based analysis. Thus, although several host traits were statistically associated with microbiome composition, each contributed only a minimal effect relative to the much larger background of inter-individual variability.

To determine whether obesity-related structure might nevertheless emerge at the taxonomic level, we next performed clustering analysis of species-level profiles (**Methods**). These clusters, which exhibit enterotype-like features, were dominated by common gut commensals, including *Bacteroides uniformis, Faecalibacterium prausnitzii, Prevotella copri, Bifidobacterium adolescentis,* and *Phocaeicola* species, and differed primarily in the relative abundance of these prevalent taxa rather than BMI category (**Fig. 1A-C**, **Supplementary Fig. 2**). Species-level abundance profiles shifted gradually across clusters, supporting a continuum of microbiome configurations rather than sharply discrete enterotypes (**Supplementary Fig. 3**). Together, these results suggest that while reproducible community structure is present in this population, it does not map strongly onto obesity-related categories.

We next examined the relationship between cluster membership and obesity-related phenotypes. BMI varied modestly across the six enterotype-like clusters, with the cluster enriched for *Phocaeicola dorei* (cluster 3, median BMI 22.50) showing a lower median BMI than the cluster enriched for *Prevotella copri* (cluster 1, median BMI of 23.75; **Fig. 1D**). Only one pairwise comparison survived multiple-testing correction (cluster 1 vs cluster 3, adjusted P=0.031 after correcting for age, sex and ethnicity), and effect sizes were small and BMI distributions overlapped substantially across all clusters, indicating limited discriminatory value for cluster membership relative to obesity status. Consistent with this, the distribution of BMI categories across clusters showed no strong enrichment of obese individuals within any single cluster (χ²=16.52, *P*=0.086; **Supplementary Fig. 4**), with all clusters containing broadly similar proportions of normal-weight, pre-obese, and obese participants.

Alpha diversity metrics, including richness and Shannon diversity, also differed significantly across clusters (Wilcoxon test *P*<0.05; **Fig. 1E**), indicating that the enterotype-like groups capture reproducible ecological variation. However, these diversity differences did not align consistently with BMI, and alpha diversity stratified directly by BMI category likewise showed substantial within-group variability and limited separation across adiposity classes (**Supplementary Fig. 5**). Together, these results indicate that while the gut microbiome in this Singaporean population exhibits reproducible community-level enterotype structure dominated by common commensals, both cluster membership and global diversity exhibit weak or no associations with obesity.

### Gut microbial taxa show limited robustness as obesity markers

We next assessed taxonomic associations with obesity across three differential abundance frameworks using three phenotype definitions: normal vs obese (NO), normal vs pre-obese (NP), and BMI as a continuous trait (Cont), and after accounting for potential confounders including age, sex, and ethnicity (**Methods**). For normal vs obese, the number of significant species varied substantially by method, with ANCOM-BC2 identifying many more associations than DESeq2 or MaAsLin2 (49 vs 1 vs 3, out of the 273 species tested; **Fig. 2A**). Despite this, only 1 overlapping species was found across two tools (ANCOM-BC2 and MaAsLin2), and no overlaps were found across all three tools (**Fig. 2A**). Similar results were observed for the other comparisons (NP and Cont), with consistently limited concordance in associations reported across methods (**Supplementary Fig. 6**). Even when multiple-testing correction was avoided and we set a threshold on nominal p-values (<0.05), no species were found consistently across all three tools and none of the observed overlaps were statistically significant (Fisher’s exact test *P*>0.05; **Supplementary Fig. 6**). Similar findings were obtained for genus and family-level association analysis, where only one taxon (Selenomonadaceae) was detected in the overlap of two tools (**Supplementary Fig. 7**). These patterns suggest that, although some obesity-related taxonomic differences can be detected, the overall signal is not robust to model choice.

**Figure 2.**
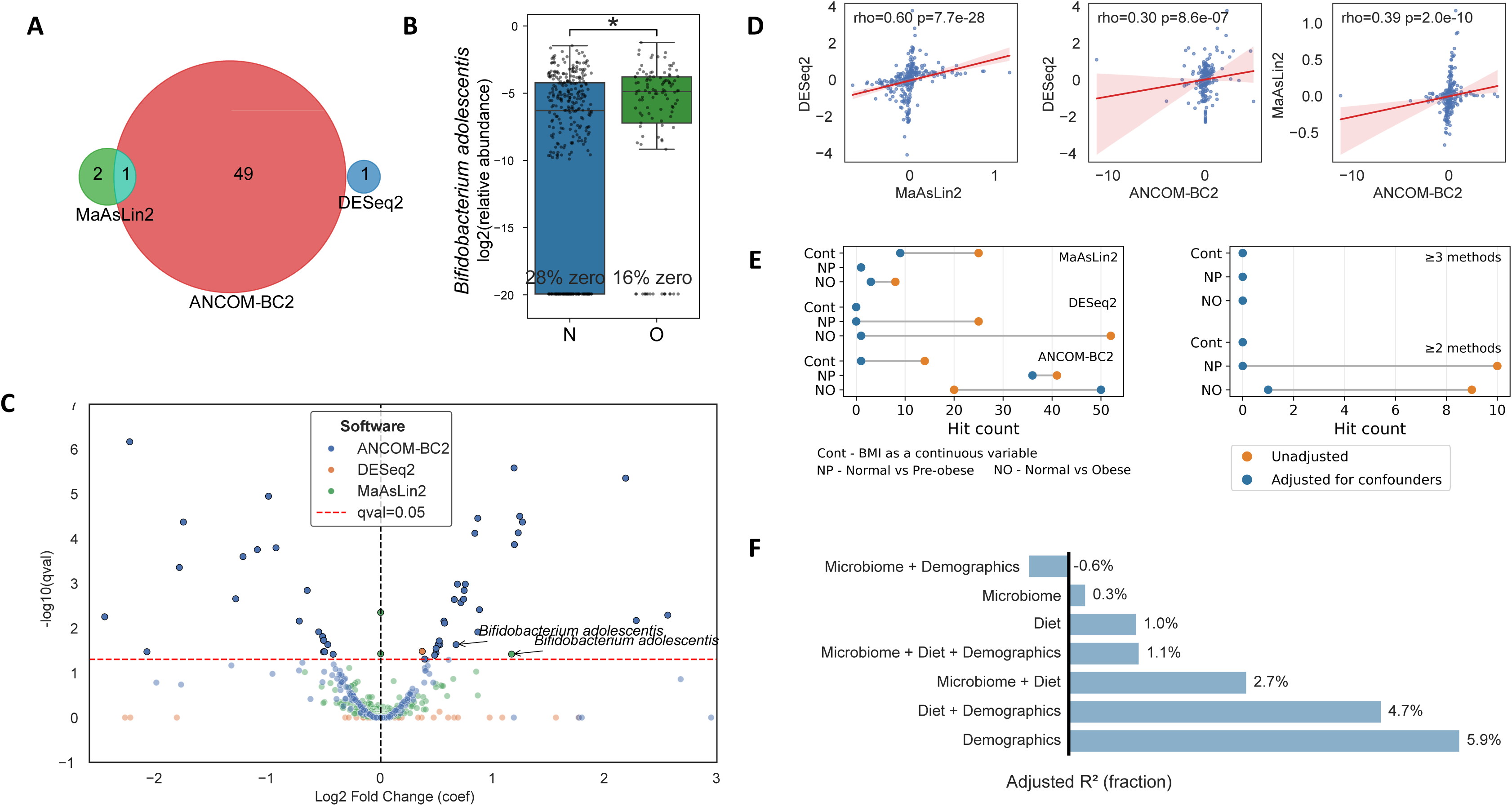
Differential taxonomic abundance analyses reveal weak, method-dependent microbiome associations with obesity. **(A)** Overlap of taxa identified as differentially abundant between normal and obese individuals (q<0.05) using three commonly applied frameworks: DESeq2, ANCOM-BC2, and MaAsLin2. Overlap across methods is limited, with no taxon identified as significant by all three approaches, indicating that the number of detected associations depends strongly on the statistical framework used. **(B)** Example taxon (*Bifidobacterium adolescentis*) illustrating a modest abundance difference between normal (N) and obese (O) individuals. Boxplots show log2-transformed relative abundance, with points representing individual samples. Substantial zero inflation is observed in both groups (28% in normal; 16% in obese), and the group distributions overlap despite statistical significance. **(C)** Volcano-style summary showing estimated effect size versus **-** log10(q) for taxa tested in the normal vs obese comparison across methods. Most taxa cluster near small effect sizes and near-threshold significance, with relatively few strong outliers. The dashed line indicates the adjusted significance threshold (q=0.05). **(D)** Pairwise comparisons of estimated effect sizes (log₂ fold change) between methods. Significant positive correlations are observed across all method pairs (Spearman’s ρ=0.30-0.60; all P<10⁻⁶), indicating modest consistency in estimated effects despite limited overlap in statistically significant taxa. **(E)** Sensitivity of differential abundance results to phenotype definition and confounder adjustment. Hit counts (q<0.05) are shown across three BMI contrasts (Cont, BMI as a continuous trait; NP, normal vs pre-obese; NO, normal vs obese) for unadjusted and confounder-adjusted models (left), together with the number of taxa detected by 2 or 3 methods (right). Although the NO contrast yielded more associations in some settings, overlap across methods remained limited, particularly after confounder adjustment. **(F)** Variation partitioning of BMI-related variation across microbiome, dietary, and demographic feature blocks, shown as adjusted R² fractions. Microbiome features alone explain only a small fraction of variation, whereas demographic variables account for the largest share, and adding microbiome features provides little additional explanatory power.

To illustrate this, we noted that while the only taxa to be detected by two tools (*Bifidobacterium adolescentis*) showed a 2-fold relative abundance change between normal and obese groups, it exhibited a wide variation in relative abundances in the normal-weight population, and a slightly higher rate at which it is not detected in this group (28% vs 16%; **Fig. 2B**). Correspondingly, the q-values assigned by both ANCOM-BC2 and MaAsLin2 for *B. adolescentis* are very close to the significance threshold (0.05), with only ANCOM-BC2 identifying a few taxa with strongly significant q-values (<0.001), but generally modest effect sizes (fold-change<2; **Fig. 2C**). Pairwise comparison of effect sizes across methods showed positive but only moderate concordance, indicating that while the direction of association was consistent, the statistical models did not agree well on the effect size (**Fig. 2D**).

To test whether the agreement exceeded chance, we applied Fisher’s exact test to the cross-method overlap: among the 274 species tested by both MaAsLin2 and ANCOM-BC2, the single species significant in both (*B. adolescentis*) did not exceed the 0.55 co-occurrences expected under independence (Fisher’s exact P=0.46). Applying the identical categorical analysis to waist circumference, WHR, and WHtR reproduced this pattern: in each case exactly one species was shared between MaAsLin2 and ANCOM-BC2, *B. adolescentis* (waist circumference, WHtR) or *A. intestini* (WHR), and in each case it failed the enrichment test (Fisher’s exact P=0.45, 0.31, 0.44). No species was shared by all three methods, and DESeq2 recovered no significant species for any measure. The recurring nominal hit was not even consistent across metrics, reinforcing that these reflect incidental method-level coincidence or borderline associations rather than strong and reproducible, obesity-specific associations (**Supplementary Table 1**).

We next assessed the impact of relaxing assumptions on our results, where as expected, in most cases adjustment for confounders notably reduced the number of significant associations detected (**Fig. 2E**). In contrast, for the normal vs obese comparison with ANCOM-BC2, substantially more taxa were reported with rather than without confounder adjustment (50 vs 20), suggesting that model specification, covariate structure, or compositional effects may influence the ANCOM-BC2 results in this contrast. Even without accounting for confounders, overlap across methods remained limited, highlighting the pitfalls of such association analysis without confounder adjustment and/or without considering the robustness of the detected associations across statistical frameworks. Similarly, relaxing the relative abundance filter (that is usually used to account for false positives in taxonomic analysis of rare taxa; **Methods**) by an order of magnitude (0.1% to 0.01%) moderately increased the number of taxa reported, without increasing cross-method overlap (**Supplementary Fig. 8**). Where overlap was observed, it was restricted to the same taxa as for the stringent threshold rather than an expanded set of concordant associations, indicating that additional hits reported by each individual method were likely driven by statistical differences near the detection limit rather than by stable biological signals. Consistent with this weak overall signal, variation partitioning showed that microbiome features explained only a small fraction of BMI-related variation at the cohort level (0.3%), though its utility was more promising in combination with dietary information (2.7%; **Fig. 2F**). Collectively, these analyses show that the number of detected microbiome associations at the taxonomic level depends strongly on analytical choices, particularly taxon filtering and the statistical framework used, and that few associations are robust across methods.

### Obesity classification with taxonomic markers achieves near-random performance

We first asked whether species-level gut microbiome composition was sufficient to classify participants as normal, pre-obese, or obese. Across elastic-net logistic regression, linear SVM, and random forest classifiers, cross-validated performance was weak, with per-class discrimination close to chance (**Fig. 3A–B**). This indicated limited generalizable BMI signal in the taxonomic profiles. Per-class AUC-ROC values clustered near the chance baseline (0.5), with only modest separation in the random forest and little evidence of meaningful discrimination by the linear models (**Fig. 3A**). The pre-obese class was especially poorly resolved across all classifiers. AUC-PR showed a similar pattern, tracking class prevalence more closely than indicating strong enrichment of true positive predictions (**Fig. 3B**). Thus, although some AUC values were slightly above chance, the absolute magnitude of discrimination was small.

**Figure 3.**
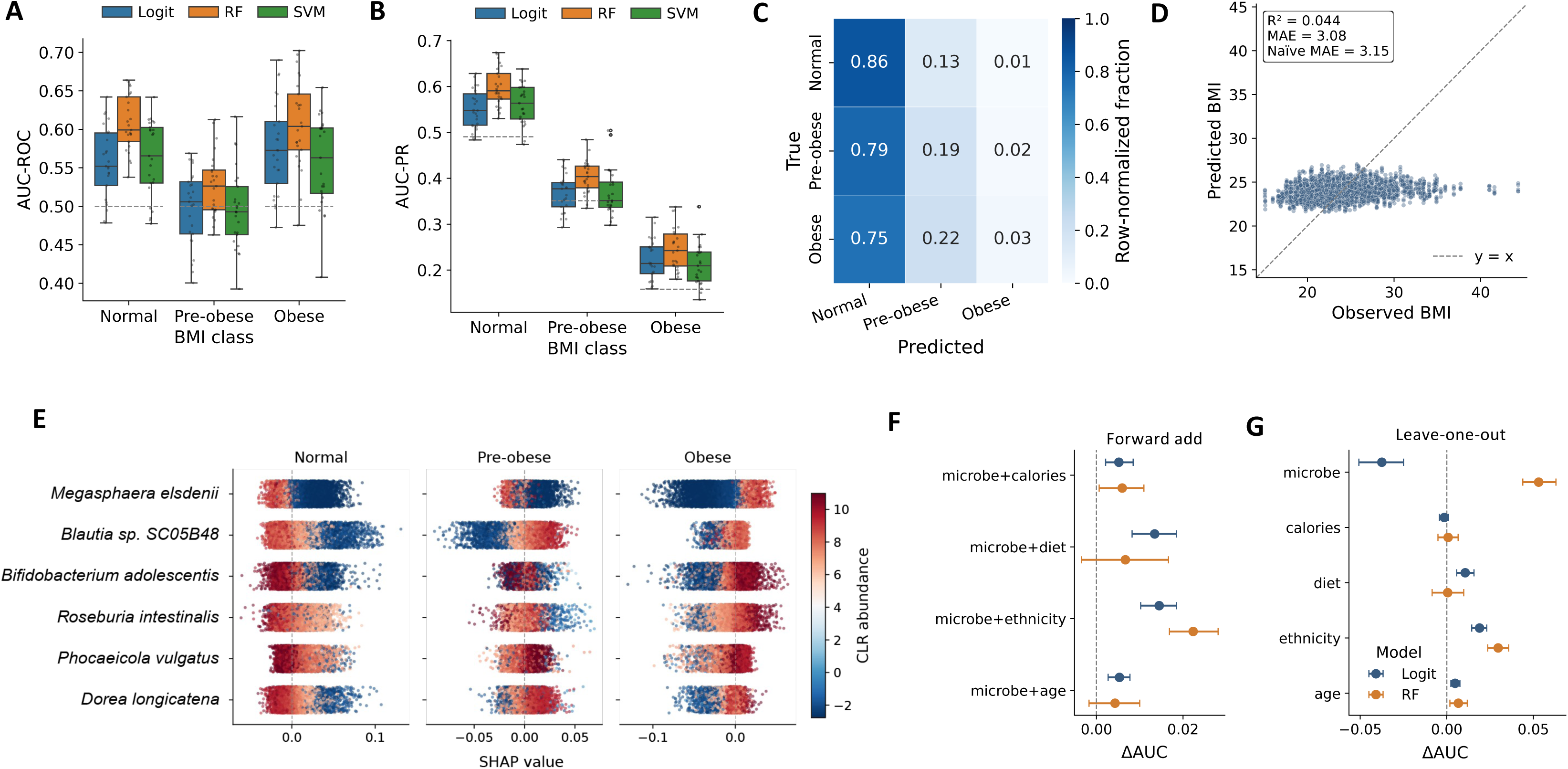
Supervised machine learning confirms limited predictive value of gut microbiome taxonomic features for obesity. **(A)** Per-class area under the ROC curve (AUC-ROC) for three classifier families — elastic-net logistic regression (Logit), random forest (RF), and linear-kernel support vector machine (SVM), predicting 3-class BMI (normal / pre-obese / obese) from CLR-transformed species abundances. Boxplots summarize the per-class AUC distribution across 25 fold-fits (5-fold stratified cross-validation, 5 repeats); dashed line marks the chance baseline (AUC=0.5). Recursive feature elimination (RFE) selected 50 features per fold. **(B)** Per-class AUC of the precision-recall curve (AUC-PR) for the same models; dashed lines mark class prevalence baselines. **(C)** Mean fold-normalized confusion matrix for the random forest base model; cells are row-normalized (rows = true class, columns = predicted class), averaged across the 25-fold. **(D)** Predicted versus observed BMI from a random forest regressor trained on continuous BMI; aggregated across 5-fold cross-validation with 5 repeats (25 folds). Dashed line: y = x. Inset reports R², mean absolute error (MAE), and the naïve cohort-mean baseline MAE. **(E)** TreeSHAP beeswarm summaries for the six features retained by RFE in all 50-fold fits of the RF model (100% selection stability). Each dot represents one held-out sample × feature; horizontal position is the SHAP value (impact on the predicted probability for the indicated class), colors indicate the sample’s CLR abundance (red = high). Peak |SHAP| ≈ 0.1 probability units i.e. even the strongest features shift the predicted class probability by at most ∼10 percentage points per sample, consistent with the near-chance AUC in panel A. **(F)** Forward-add forest plot: ΔAUC = AUC(microbe + block) − AUC(microbe-only), paired per fold; 95% bootstrap CI across 25 paired fold-differences. Positive values indicate the block adds information beyond the microbiome. **(G)** Leave-one-out forest plot: ΔAUC = AUC(joint model) − AUC(joint model without block), paired per fold; the joint model includes microbiome + calories + diet + ethnicity + age. Positive values indicate the block uniquely contributes information beyond all others.

Permutation-label testing confirmed that most models were indistinguishable from the shuffled-label null (**Supplementary Fig. 9**). Logistic regression (logit) did not reach statistical significance for any BMI class (normal p = 0.10, pre-obese p = 0.30, obese p = 0.38). Random forest (RF) achieved significance for the normal and obese classes (AUC = 0.596, p = 0.001 and AUC = 0.599, p = 0.010, respectively), but not for pre-obese (AUC = 0.522, p = 0.21), indicating that the only reproducible signal was confined to the two BMI extremes and required a non-linear model. Importantly, the significant RF effects remained quantitatively small: the observed AUCs were approximately 0.60 for normal and obese, far below values that would indicate practical classification utility. The same negative outcome held for a classifier trained to distinguish at-risk central adiposity (waist circumference, WHR and WHtR) using an identical pipeline, where it failed to exceed chance across all three families, reproducing the near-chance BMI result (**Supplementary Table 1**). These results support the presence of a weak non-linear microbiome signal, but not a robust predictive classifier.

The confusion matrices further showed that above-chance ranking did not translate into useful class assignment. The random forest largely collapsed toward normal predictions, correctly classifying most normal samples but very few pre-obese or obese samples (**Fig. 3C**). In contrast, the linear models distributed predictions diffusely across all three classes, with diagonal accuracies near the one-third chance expectation - logistic regression: 0.43 / 0.35 / 0.37 (normal / pre-obese / obese); linear SVM: 0.44 / 0.33 / 0.33 (**Supplementary Fig. 10**). The off-diagonal errors were not concentrated between adjacent BMI categories, arguing against a simple “noisy ordinal” model in which the classifiers captured BMI ordering but failed at exact class boundaries. Instead, the models failed in different ways while converging on near-chance categorical accuracy.

The negative classification result was not an artefact of discretizing BMI into classes. Continuous BMI regression produced the same conclusion. Random-forest regression explained only a very small fraction of BMI variance, with R² = 0.044 and MAE = 3.08 BMI units, only marginally better than a naïve cohort-mean predictor with MAE = 3.15 (**Fig. 3D**). The prediction cloud was compressed around the cohort mean rather than tracking observed BMI across its range. ElasticNet regression performed worse than the naïve baseline, with negative R². Thus, whether BMI was treated as categorical or continuous, microbiome taxonomic features provided little generalizable predictive information.

Feature attribution analyses were consistent with a weak global gradient rather than discrete BMI-specific microbial signatures. In the random forest, a small set of common taxa was selected reproducibly across folds, including *Megasphaera elsdenii*, *Bifidobacterium adolescentis*, *Roseburia intestinalis*, *Phocaeicola vulgatus*, *Blautia* sp. SC05B48, and *Dorea longicatena* (**Fig. 3E**). TreeSHAP values indicated that these taxa contributed modestly to class probabilities, with the strongest individual effects shifting predicted probabilities by only about 0.1. The same features tended to move predictions along a normal-versus-obese axis, while the pre-obese class showed little distinct attribution pattern. This supports the interpretation that the random forest captured a small compositional gradient, not a set of strong class-specific biomarkers.

To quantify the microbiome’s contribution relative to host factors, predictor blocks were compared in two complementary ways; in both, a positive ΔAUC indicates that the block adds predictive information. The first, forward-add analysis began from a microbiome-only model and added each host block in turn (**Fig. 3F**). Demographic and dietary blocks produced small but detectable gains, confirming that the pipeline could recover predictive signal when present and that the microbiome-only baseline was weak. The second, leave-one-out analysis began from the full model (microbiome + calories + diet + ethnicity + age) and removed each block to isolate its unique contribution (**Fig. 3G**). Here the microbiome’s contribution was strongly model-dependent: in the linear models its removal slightly improved AUC (negative ΔAUC), so the high-dimensional taxonomic matrix acted mainly as noise under a linear decision boundary; in the random forest its removal reduced AUC (positive ΔAUC), indicating a small, unique non-linear contribution. Even in this most favorable setting, the microbiome’s contribution was modest and comparable to individual host-covariate blocks.

Taken together, these analyses argue against strong BMI predictability from species-level gut microbiome composition in this cohort. Linear classifiers showed no statistically significant per-class signal, continuous linear regression underperformed a naïve baseline, and categorical predictions were near chance. A random forest detected a statistically significant but small signal for the normal and obese extremes, supported by reproducible but low-magnitude SHAP attributions. However, this signal did not yield clinically useful discrimination, did not resolve the pre-obese class, and explained little continuous BMI variation. Thus, the microbiome profile contains at most a weak non-linear correlate of BMI status, rather than a robust predictive signature

### Functional profiling reveals sparse, method-dependent metabolic associations with obesity

To assess whether obesity is associated with coordinated functional shifts in the gut microbiome, we analyzed MetaCyc pathway profiles derived from shotgun metagenomes while adjusting for age, sex, and ethnicity. Ordination of pathway-level profiles using Bray-Curtis dissimilarity showed broad dispersion of samples within BMI categories, with no clear separation by BMI category or ethnicity (**Fig. 4A**). This pattern closely mirrored taxonomic ordination results (**Fig. 1A**, **Supplementary Fig. 1**), suggesting that functional potential, like taxonomic composition, is dominated by other sources of individual-specific variation rather than obesity-related structure. Consistent with this observation, PERMANOVA indicated that BMI explained only a small fraction of functional variation (R² = 0.0046, P = 0.001), weaker than that observed for taxonomic profiles, indicating that global functional reprogramming is not a dominant feature of obesity in this cohort.

**Figure 4.**
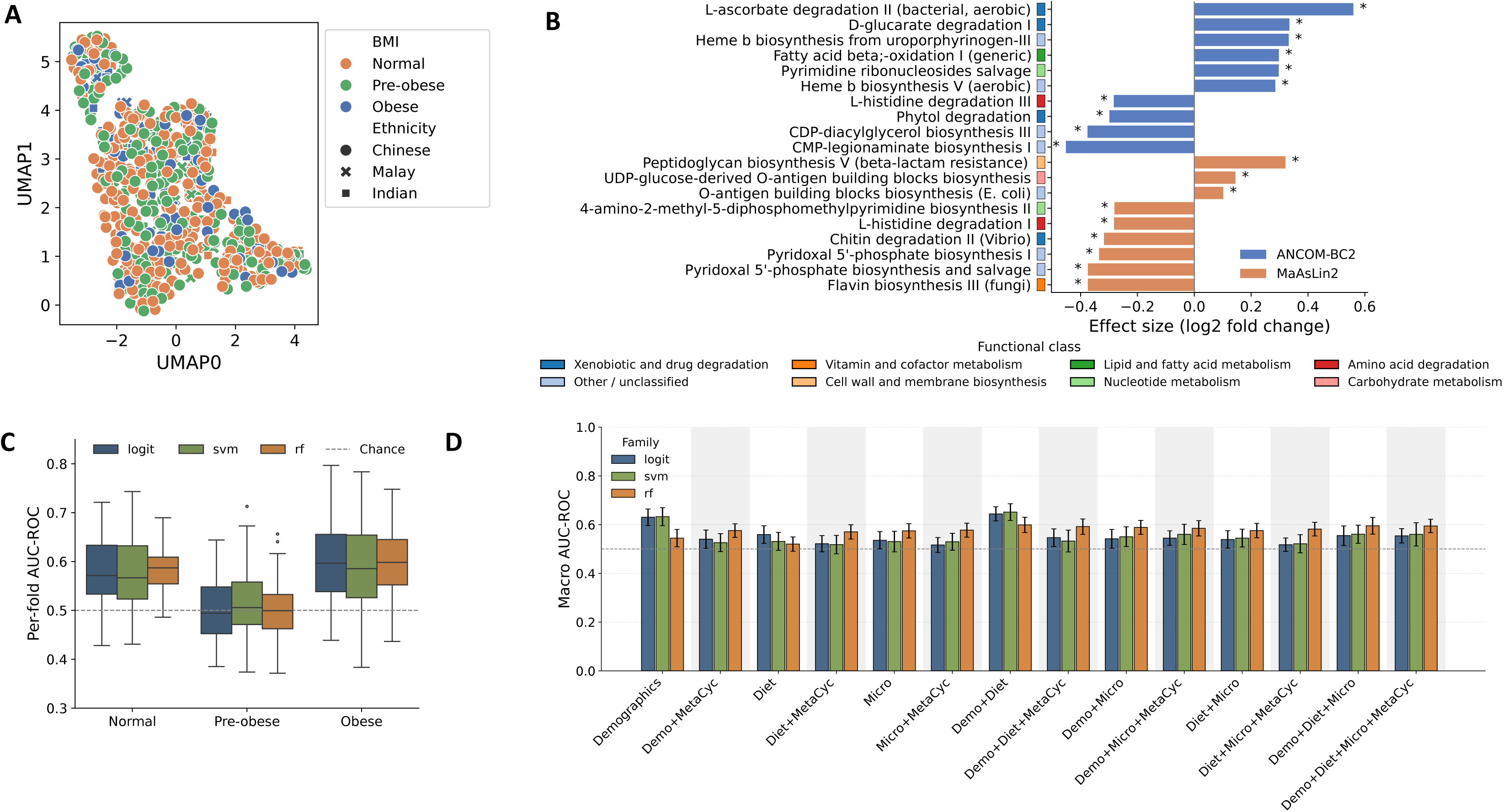
MetaCyc functional pathway profiles are not associated with BMI. **(A)** UMAP projection of Bray-Curtis dissimilarities computed from MetaCyc functional pathway relative abundances. Each point represents one sample, colored by BMI class (normal = orange, pre-obese = green, obese = blue) and with shapes indicating ethnicity (Chinese = circle, Malay = ×, Indian = square). BMI classes are intermixed throughout the UMAP space with no discernible clustering, indicating that overall functional community structure does not stratify by obesity status. **(B)** Horizontal forest plot of differential abundance effect sizes (log₂ fold change, obese vs normal) from ANCOM-BC2 (blue) and MaAsLin2 (orange), adjusted for age, sex, and ethnicity. Bars represent pathways with the largest absolute effect sizes; asterisks denote q < 0.05 after FDR correction. Bar color indicates functional class. Despite a number of nominally altered pathways, inter-method concordance is poor (see **Supplementary Figure 11**), and hits span diverse, biologically unrelated functional classes. **(C)** Per-fold one-vs-rest AUC-ROC from 5×5 stratified cross-validation with repeats (25 folds) for predicting each BMI class from MetaCyc relative abundances, across three classifier families: Elastic-Net Logistic Regression (logit, blue), Linear SVM (green), and Random Forest (orange). AUC is near chance across all classes and classifiers, with pre-obese showing the lowest discrimination. Performance does not exceed chance after permutation testing (**Supplementary Figure 13**). **(D)** Macro-averaged AUC-ROC (mean ± SD across folds) for all 14 feature-set combinations spanning the three data modalities - demographics (demo), diet, and microbiome (Micro) - each tested with and without MetaCyc pathway features (grey shading), across three classifier families. Demographics-only achieves the highest macro AUC (logit=0.62), reflecting signal from age, sex, and ethnicity. Adding MetaCyc to any feature combination does not consistently improve performance, and MetaCyc alone performs at or near chance for all classifiers, confirming that functional pathway profiles carry no independent discriminative information for BMI category in this cohort.

We next performed differential abundance analysis for MetaCyc pathways comparing normal and obese individuals using ANCOM-BC2 and MaAsLin2 (DESeq2 requires count data). Pathway-level associations were sparse, with most effect sizes centered near zero (**Fig. 4B**). Only a limited number of pathways passed multiple-testing correction, and concordance between the two methods was poor. At the level of individual pathways, there was no overlap between method-significant hits at q < 0.05: all significant associations were method-specific (**Supplementary Fig. 11**). When pathways were aggregated by functional category, partial thematic overlap was apparent, for example, distinct histidine degradation variants (Histidine degradation I, Histidine degradation III) were each flagged by one method, but this reflected convergence on a broader metabolic class rather than agreement on specific pathways. No higher-order aggregation by metabolic category emerged with consistent directionality or effect size across both methods, further supporting the absence of coherent functional reprogramming associated with obesity.

Consistent with these weak pathway-level signals, machine learning models trained on MetaCyc pathway abundances alone achieved per-class AUROC values only marginally above chance across all three BMI classes and classifier families (**Fig. 4C; Supplementary Fig. 12**). Permutation testing confirmed that observed above-chance values were not statistically significant (**Supplementary Fig. 13**). Normalised confusion matrices revealed a strong tendency to misclassify pre-obese samples as normal, reflecting the overlapping phenotypic boundary between these classes (**Supplementary Fig. 14**). TreeSHAP analysis of the Random Forest classifier identified a small set of pathways as primary drivers of classification (**Supplementary Fig. 15**); however, mean absolute SHAP values were negligible across all features (maximum = 0.013, median = 0.005), corresponding to shifts of less than 2 percentage points in predicted class probability. The low SHAP value quantitatively confirms that the classifier was operating near random, consistent with the weak differential signal observed in formal testing.

To assess the incremental value of functional features in a multi-modal context, we evaluated macro-averaged AUROC across all 14 feature-set combinations spanning demographics, diet, and microbiome, each tested with and without MetaCyc pathway features across three classifier families (**Fig. 4D**). Demographics-only classifiers achieved the highest macro AUROC (logit ≈ 0.62), reflecting signal from age, sex, and ethnicity. Adding MetaCyc features to any combination did not consistently improve performance across classifier families, and microbiome alone performed near chance. A notable exception was Random Forest: adding MetaCyc features to single-modality feature sets yielded modest gains in macro AUC for RF specifically (demographics: +0.031; diet: +0.050), suggesting that the ensemble structure of RF can extract weak non-linear signal from high-dimensional pathway data that linear classifiers cannot. However, this advantage did not persist as additional modalities were combined. RF with the full feature set (demographics+diet+microbiome+MetaCyc, AUC=0.594) showed negligible improvement over demographics+diet+microbiome without MetaCyc (AUC=0.595), and RF macro AUC remained well below the best-performing linear models on demographics alone (logistic regression and SVM on demographic+diet: AUC ≈ 0.64–0.65). The RF-specific gains from MetaCyc may therefore reflect the susceptibility of tree ensembles to spurious correlations in high-dimensional data rather than genuine discriminative signal. Together, these results show that functional metagenomic profiling reinforces the conclusions drawn from taxonomic analyses: obesity-associated microbiome signals in this cohort are weak and diffuse, rather than reflecting a strong biologically meaningful functional signature.

## Discussion

In this study, we conducted a population-scale analysis of gut microbiome composition, functional potential, and predictive modeling in relation to obesity in a multi-ethnic Asian cohort living within a shared urban environment. By combining deep shotgun metagenomic profiling with standardized demographic and anthropometric measurements from 871 HELIOS participants, including ethnic Chinese, Malay, and Indian adults, this study provides one of the largest metagenomic assessments of obesity-related microbiome variation in Southeast Asia. Across ecological, taxonomic, functional, and machine-learning-based analyses, we found few strong or reproducible microbiome signals associated with obesity. While differences in enterotype-like structure, differentially abundant taxa, and pathway-level differences were detectable, effect sizes were modest, concordance across analytical frameworks limited, and the corresponding signals did not support robust discrimination of obesity status at the population level. Variance partitioning quantified the magnitude of this signal directly: microbiome features collectively explained only 0.3% of BMI variance, compared with 5.9% for demographic factors and 1.0% for diet (**Fig. 2F**). Together, these findings suggest that in this setting, the gut microbiome likely explains obesity-related variation in the population to a very limited extent, especially when compared with host demographic and dietary factors.

Our results refine and contextualize earlier reports linking gut microbiome features to obesity. Previous large-scale human cohort studies have described differences in microbial diversity, taxonomic composition, and functional profiles between individuals with obesity and lean controls, often by combining data across geographically, culturally, or clinically distinct populations^6,7,28^. Such findings may reflect real microbiome signals, but those signals may also be context-dependent and more apparent in studies comparing more divergent environments, more extreme phenotypes, or cohorts with greater heterogeneity in diet and lifestyle. Subsequent meta-analyses, however, have shown that these signals are inconsistent across cohorts and that no single taxon-level marker reliably distinguishes obesity status across studies^13,14^. Our findings establish that this emerging consensus is indeed valid in a Southeast Asian population. By analyzing a large urban cohort that shares healthcare, food supply, and infrastructure within a single city-state, our study minimizes the environmental heterogeneity that typically inflates between-cohort comparisons and provides a stringent test for population-level obesity-microbiome signatures. Under these conditions, we observed broad overlap in global microbiome structure across BMI groups and only a few recurring taxonomic and pathway-level signals, each characterized by modest effect size, overlapping group distributions, and limited consistency across univariate, multivariate, and predictive analyses. Our findings also align with the ongoing reassessment of early obesity–microbiome markers. The *Firmicutes:Bacteroidetes* ratio and reduced α-diversity, proposed as hallmarks of obesity in foundational studies, showed no association with BMI in this cohort, whether modelled continuously or across BMI categories (Spearman ρ≤0.06, P ≥ 0.13; Kruskal – Wallis P ≥ 0.21; **Supplementary Figure 16**), consistent with subsequent large-scale analyses that have failed to reproduce these signals. These findings should therefore be seen as evidence that many previously reported associations may be cohort-sensitive, impacted by confounders, or difficult to generalize at population scale.

The limited overlap across differential abundance methods likely reflects the underlying structure of the data rather than a simple distinction between “better” and “worse” tools, a pattern previously documented across diverse microbiome datasets^25,29^. ANCOM-BC2 identified more associations than MaAsLin2 and DESeq2 under several contrasts, likely reflecting its bias-corrected log-linear modelling of sampling fractions, which can be more sensitive than negative-binomial or normalize-then-fit alternatives in sparse, low-effect settings. A particularly clear indicator of this sensitivity was the ANCOM-BC2 result for the normal versus obese contrast, where covariate adjustment increased rather than decreased the number of significant hits (50 vs 20), contrary to the expectation that confounder adjustment would generally reduce the number of associations, suggesting sensitivity to model specification, covariate structure, or compositional effects rather than a stable biological signal. By contrast, pairwise effect-size correlations between methods were positive (ρ = 0.30–0.60; **Fig. 2D**), indicating that methods broadly agreed on the direction of association even when they disagreed on which features crossed the significance threshold. The resulting low cross-method overlap suggests that many obesity-associated signals in this cohort lie close to the detection boundary rather than representing strong, cohort-wide effects that can be seen as biological signals. This interpretation is further supported by two additional observations: relaxing abundance filtering increased the number of detected hits without meaningfully increasing cross-method overlap, and the same pattern persisted at coarser taxonomic resolution, with only the family Selenomonadaceae appearing in any two-method overlap at the genus/family level (**Supplementary Fig. 7**).

Three independent lines of evidence in our analyses converge on a common interpretation: any microbiome signal in this cohort is non-linear and concentrated at the extremes of the BMI distribution rather than graded across its full range. First, categorical BMI contrasts yielded substantially more differentially abundant features than continuous BMI analyses across all three methods (**Fig. 2E**), consistent with categorization enriching for contrasts at the ends of the adiposity spectrum while continuous modelling diluting such effects through within-category heterogeneity and measurement noise. Second, the divergence between linear and non-linear classifiers was striking: neither logistic regression nor linear SVM achieved per-class AUC significantly above the permutation null (all p > 0.10), whereas the random forest detected weak but statistically significant discrimination for the Normal class (AUC = 0.596, p = 0.001) and the Obese class (AUC = 0.599, p = 0.01) but not for the Pre-obese class (AUC = 0.522, p = 0.21; **Supplementary Fig. 9**). The ability of a non-linear model to detect weak signal where linear models did not is consistent with a non-linear or interaction-dependent microbiome-BMI association^30^. Third, the pre-obese class was the consistent point of failure across every analytical framework (differential abundance, classification, regression) with classifiers preferentially misclassifying pre-obese samples toward normal rather than toward obese (**Fig. 3C; Supplementary Fig. 10**). This may help explain why normal versus obese contrasts typically yielded more associations than continuous BMI analyses in our dataset. Together, these observations suggest the microbiome may shift in association with adiposity only when adiposity crosses some threshold, rather than tracking BMI smoothly. Such threshold-like behavior would be consistent with ecological shifts in gut community composition, in which reorganization may occur around phenotype boundaries rather than scaling continuously with host BMI.

The few recurring taxonomic signals are likewise better viewed as candidate context-dependent markers than as universal obesity microbiome signatures. *Bifidobacterium adolescentis* and the family Selenomonadaceae emerged across univariate analyses as the more reproducible signals, though both sat close to the multiple-testing threshold (q ≈ 0.05) and *B. adolescentis* exhibited substantial zero inflation (28% Normal vs 16% Obese; **Fig. 2B**). Both are common healthy gut commensals rather than canonical obesity-specific markers; *B. adolescentis* is a widely distributed fibre and starch fermenter^31^, and their recurrence here is more consistent with chance overlap between methods (Fisher’s exact *P*>0.05) than with a direct, obesity-specific role. Variance partitioning supports this: microbiome and dietary features each carried partly independent information about BMI (combined 2.7% vs 0.3% for microbiome alone and 1.0% for diet alone; **Fig. 2F**), indicating that any apparent microbiome-BMI association cannot be cleanly separated from dietary context. The random forest feature-attribution analysis (**Fig. 3E**) identified a partially distinct set of taxa, including *Megasphaera elsdenii, Blautia sp.* SC05B48*, Roseburia intestinalis, Phocaeicola vulgatus,* and *Dorea longicatena*, with only *B. adolescentis* appearing in both the univariate and multivariate analyses. This represents a feature-level discordance distinct from the model-family divergence noted above: univariate differential abundance methods disagreed with each other on which features were significant, and the features prioritized by multivariate machine learning were largely different again. Notably, none of these taxa correspond to the canonical obesity-associated markers reported in earlier Western cohorts^6,7,28^, further arguing against transferable population-level signatures. In all cases, effect sizes were small (fold-change < 2; **Fig. 2C**), abundance distributions overlapped extensively across BMI groups, zero inflation was substantial, and SHAP magnitudes confirmed that even the most influential individual taxa shifted predicted class probability by ≤ 0.1 (**Fig. 3E**). Together, these observations argue that no single taxon emerges as a reproducible, framework-independent marker of obesity in this cohort, even at the extremes of BMI where any signal is concentrated.

The pathway-level results point in a similar direction. Functional ordination showed little separation by BMI category, BMI explained only a small fraction of total pathway variation (R² = 0.0046, P = 0.001), and pathway-based machine-learning classification achieved per-class AUC only marginally above chance across all three classifier families (**Fig. 4C**). Critically, adding MetaCyc features to any combination of demographic, dietary, and taxonomic features did not consistently improve classification performance over demographics alone (**Fig. 4D**), indicating that functional metagenomic profiles add little incremental discriminative information for BMI beyond host covariates. At the individual pathway level, there was no overlap between method-significant hits at q < 0.05; when pathways were aggregated by functional category, partial thematic overlap emerged (distinct histidine degradation pathways were each flagged by one method) but this reflected convergence on a broader metabolic class rather than agreement on specific pathways. Together, these observations suggest that obesity-related functional differences in this cohort are subtle, sparsely distributed, and not organized into a coordinated metabolic program.

A related possibility is that biologically meaningful differences reside below the taxonomic and functional resolution captured by short-read metagenomic profiling. Closely related strains can differ markedly in gene content, substrate utilization, and metabolite production^32^, and pathway annotations from reference databases capture inferred catalytic potential rather than active expression. Weak species- and pathway-level signals therefore do not necessarily imply that the microbiome is biologically irrelevant to obesity; the relevant differences may instead be strain-resolved, pangenomic, or expressed only under specific dietary or physiological conditions. Future studies integrating long-read or strain-aware metagenomic profiling, metatranscriptomics, and stool or serum metabolomics will be necessary to determine whether obesity-associated microbial differences are being masked by the resolution limits of conventional shotgun analysis.

Several considerations delimit the scope and interpretation of our findings. First, this study is cross-sectional in design, using baseline stool samples from a prospective cohort; we can therefore infer association but not direction of causation between microbiome composition and adiposity. Second, residual heterogeneity in diet, medication exposure, socioeconomic factors, and lifestyle may obscure associations present only within narrower strata. To probe this, we repeated the differential-abundance analysis within the two subgroups best powered in this cohort (ethnicity and sex) each adjusted for the remaining demographic covariates, including age as a continuous term. The null persisted in every stratum: the conservative method (MaAsLin2) recovered at most one species, and no taxon was reproducible across methods beyond chance (Fisher’s exact P ≥ 0.18, **Supplementary Table 2**). It nonetheless remains plausible that associations are more detectable within finer or differently defined strata, dietary patterns or metabolically unhealthy obesity, that we were not powered to resolve. Third, obesity is a heterogeneous phenotype. Although BMI is standard for epidemiological comparability, our null results were robust to the choice of anthropometric adiposity metric. Waist circumference, WHR, and WHtR each reproduced the near-null compositional variance, yielded no differential-abundance associations that replicated across methods, and supported only near-chance classification; where a taxon recurred (*B. adolescentis*), it did so as a nominal, single-method signal that failed cross-method and enrichment testing (**Supplementary Table 1**). Deeper adiposity phenotypes such as DEXA-derived body composition, HOMA-IR-based insulin resistance, liver fat fraction, and inflammatory or cardiometabolic biomarkers, may reveal stronger or qualitatively different microbiome associations. Finally, because this study was conducted in an Asian cohort in Singapore, the results may not directly generalize to populations with different environmental, dietary, or genetic backgrounds, though they extend the obesity-microbiome literature to a previously underrepresented Southeast Asian population.

Taken together, the consistency of weak signals across taxonomic, functional, and predictive analyses argues against a strong, broadly generalizable gut microbiome signature of obesity in this cohort. Microbiome composition explained substantially less BMI variance than demographic or dietary factors (0.3% vs 5.9% and 1.0%, respectively; **Fig. 2F**), and where microbiome signals were detectable they were small in magnitude, concentrated at the BMI extremes, and not robust across analytical frameworks. Our findings therefore support a model in which the microbiome contributes alongside stronger host and environmental determinants of adiposity, and caution against framing obesity as a condition in which the gut microbiome plays a dominant population-level role. Resolving whether any reproducible microbiome correlates of obesity exist in human populations will likely require strain-resolved metagenomic profiling, and integration of paired metatranscriptomic and metabolomic data, perhaps in larger and more deeply phenotyped cohorts.

## Methods

### Study design and participants

This study used baseline stool samples from participants enrolled in the Health for Life in Singapore (HELIOS) study, a population-based prospective cohort initiated in 2017. The study was approved by the Nanyang Technological University Institutional Review Board (IRB-2016-11-030), and all participants provided written informed consent. Singaporean citizens or permanent residents aged 30–84 years were recruited from the general population through community-based engagement strategies designed to ensure representation across ethnic groups, socioeconomic strata, and working-age adults. Individuals who were pregnant, breastfeeding, acutely ill, or unable to provide informed consent were excluded. For the present analysis, stool samples from 871 participants of Chinese, Malay, and Indian ethnicity were included.

### Demographic and anthropometric measurements

Participant sex, age, and ethnicity were obtained from national registry records. Height and weight were measured using a BSM 370 automatic stadiometer (InBody, Seoul, South Korea). Body mass index (BMI) was calculated as weight in kilograms divided by height in metres squared. BMI categories were defined using WHO Asian-population cutoffs. Participants with BMI <23.0 kg/m², including underweight participants, were grouped as normal-weight for analysis, pre-obese if BMI was 23.0–27.4 kg/m², and obese if BMI was ≥27.5 kg/m². Waist circumference was measured at the midpoint between the lowest rib and the iliac crest, following WHO and International Diabetes Federation recommendations. For categorical analyses, each adiposity measure was dichotomised into "at-risk" versus "normal" using established clinical cut-points: waist circumference by the International Diabetes Federation (IDF) Asian thresholds for central obesity (≥90 cm in men, ≥80 cm in women); WHR by the World Health Organization thresholds for substantially increased risk (≥0.90 in men, ≥0.85 in women); and WHtR by the sex- and ethnicity-independent boundary value ≥0.5.

### Stool sample collection

Participants collected stool samples using the Fe-Col® collection device (Alpha Laboratories) together with DNA/RNA Shield™ Fecal Collection Tubes (Zymo Research). Each tube contained 9 mL of DNA/RNA Shield™, which preserves nucleic acids and inactivates microorganisms at ambient temperature. Approximately 1g of stool was collected using the spoon attached to the tube cap and mixed thoroughly with the preservative by inversion. Samples collected at home were returned by mail and subsequently stored at −80°C until DNA extraction.

### DNA extraction and shotgun metagenomic sequencing

DNA was extracted from stool samples using the QIAamp PowerFecal Pro DNA Kit (Qiagen) following the manufacturer’s protocol. Extracted DNA was eluted in 50µL of elution buffer and quantified using the QuantiFluor dsDNA System (Promega) on a VICTOR Nivo multimode microplate reader (PerkinElmer). Metagenomic libraries were prepared using the NEBNext Ultra II FS DNA Library Prep Kit for Illumina (New England Biolabs) and indexed with NEBNext Multiplex Oligos. Library fragment size and concentration were assessed using an Agilent 4200 TapeStation system. Paired-end sequencing (2×150 bp) was performed on an Illumina HiSeq X platform, yielding a mean sequencing depth of >20 million reads per sample. Three blank collection tubes were processed in parallel as negative controls.

### Taxonomic and functional profiling

Adapter trimming and quality filtering were performed using fastp^33^ (v0.20.1). Host-derived reads were removed by alignment to the human reference genome (hg19) using BWA-MEM^34^ (v0.7.17). The remaining microbial reads were used for taxonomic profiling with Kraken2^35^ (v2.0.8) and abundance estimation with Bracken^36^ (v2.5), using the standard 16GB microbial database (December 2022 release). Taxa detected at relative abundances below 0.1% were excluded to reduce sparsity and minimize false-positive calls, and profiles were renormalized after filtering.

Potential contaminant taxa were screened using DNA yield as a proxy for contamination risk, based on the expectation that reagent-derived contaminants are enriched in low-biomass samples. Within each sequencing batch, we computed the Spearman correlation between each taxon’s relative abundance and sample DNA yield. Taxa were flagged as contaminant candidates if they showed both a negative association with DNA yield and evidence of detection in negative controls. Specifically, candidate contaminants were defined as taxa with Spearman’s ρ ≤ −0.4 and mean relative abundance >0.1% in at least one negative-control sample. Potential contaminant taxa were filtered out from taxonomic profiles before renormalization.

Functional profiling was performed using HUMAnN3^37^ (v3.1.0) with default parameters and the UniRef90 database. MetaCyc pathway abundances were obtained from HUMAnN3 outputs, excluding unmapped and unintegrated pathways.

### Diversity, ordination and cluster analyses

Alpha diversity was assessed using Shannon and richness indices. Beta diversity was evaluated using Bray-Curtis dissimilarity and visualized via ordination methods including principal coordinate analysis (PCoA) and Uniform Manifold Approximation and Projection (UMAP). Differences in beta diversity were tested using permutational multivariate analysis of variance (PERMANOVA) with 9999 permutations.

To identify reproducible community configurations, spectral clustering was applied to a high-dimensional UMAP embedding of species-level relative abundance profiles. A nearest-neighbor affinity graph (n_neighbors = 80) was constructed and cluster assignments derived via k-means on the spectral embedding (eigen_solver = "arpack"). The number of clusters k was selected over k = 2–6 by combining bootstrap stability as the primary criterion with silhouette coefficient as a tiebreaker. Bootstrap stability was estimated by refitting spectral clustering on 80% subsamples across 20 replicates and computing, for each sample, the proportion of replicates in which it was co-assigned to the same cluster as its neighbors (consensus matrix approach). Per-sample stability scores were retained as a continuous measure of assignment confidence.

### Differential abundance analysis

Associations between microbial taxa or functional pathways and adiposity were assessed using a consensus differential-abundance framework comprising DESeq2^38^, ANCOM-BC2^39^, and MaAsLin2^40^. These methods were chosen because they represent complementary approaches to microbiome association testing: DESeq2 models sequencing counts using a negative-binomial framework, ANCOM-BC2 addresses compositional bias in relative-abundance data, and MaAsLin2 provides a flexible multivariable modelling framework for covariate-adjusted associations. Because benchmarking studies^29^ have shown that differential-abundance results can be sensitive to method choice, false-discovery control, and confounding structure, we used cross-method agreement to prioritize robust associations over method-specific findings. Features detected by at least two of the three methods were therefore considered the higher-confidence consensus set. Models were adjusted for age, sex, and ethnicity unless otherwise specified. Multiple testing correction was performed using the Benjamini–Hochberg procedure. Taxa or pathways identified by multiple methods were considered more robust, although all results were interpreted in the context of effect size, prevalence, and consistency across approaches. To assess whether obesity-associated differential abundance was masked at the whole-cohort level, the three-tool analysis (MaAsLin2, ANCOM-BC2, DESeq2; Obese vs Normal) was repeated within ethnicity (Chinese) and sex (female, male) subgroups, each adjusted for the remaining demographic covariates with age modelled continuously; cross-method overlap was assessed by Fisher’s exact test.

### Classification and regression models

Supervised machine-learning-based classifiers were trained to predict BMI categories using taxonomic profiles. Taxonomic abundances were normalized using centered log-ratio (CLR). A pseudocount of 1 was added prior to log-ratio transformations.

Three classifier families were evaluated to compare linear and non-linear decision boundaries. Elastic-net logistic regression was fitted using balanced class weights, an elastic-net penalty with equal L1 and L2 mixing, and internal feature standardization. Linear-kernel support vector machines were fitted with balanced class weights and internal feature standardization. A linear kernel was used to preserve compatibility with recursive feature elimination, which requires access to model coefficients. Random forest classifiers were fitted with 500 trees, square-root feature subsampling, and balanced class weights; no feature standardization was applied because random forests are scale-invariant.

### Recursive feature elimination

Recursive feature elimination was performed within each outer training fold using sklearn.feature_selection.RFE. Unless otherwise stated, models were trained with a target of 50 selected features and a step size of 50% per elimination round. The selected feature count was chosen from a feature-count sensitivity sweep showing that held-out performance plateaued once approximately 45 or more features were retained.

Feature importance for RFE was calculated as the mean absolute model coefficient for logistic regression, ElasticNet, and linear SVM models, and as Gini importance for random forest models. For analysis scenarios that included host covariate blocks, these covariates were forced into the selected feature set so that the model always retained access to the specified non-microbial predictors. In scenarios where the total number of available predictors was smaller than the target feature count, RFE was not applied and the model was trained on the full available feature set.

### Confounder analysis

To assess the contribution of microbiome features relative to host covariates, predictors were grouped into predefined blocks. The calories block contained a single feature: total daily energy intake (kcal day⁻¹). The diet block contained 12 macronutrient features - nine absolute daily intakes (total protein, total fat, saturated / monounsaturated / polyunsaturated fatty acids, total carbohydrate, starch, sugar, and dietary fibre, all in g day⁻¹) and three per-kcal densities (protein, carbohydrate, and total fat in g per 1000 kcal). The microbiome block consisted of the centered log ratio (CLR) transformed species features. The ethnicity block contained three one-hot ethnicity indicators (Chinese, Indian, Malay), and the age block contained chronological age.

Two complementary block-level analyses were performed. In the forward-add analysis, each non-microbial block was added individually to the microbiome-only model, and the resulting per-fold AUC was compared with the microbiome-only baseline. In the leave-one-out analysis, a joint model containing microbiome, calorie, diet, ethnicity, and age predictors was fitted, then refitted after removing one block at a time. Paired per-fold AUC differences were used to estimate the unique contribution of each predictor block conditional on the others.

### Permutation-label null testing

Permutation-label testing was used to assess whether classifier performance exceeded the shuffled-label null distribution. For random forest and logistic regression models, BMI labels were randomly permuted across samples and models were refitted under a single stratified 5-fold cross-validation pass. This procedure was repeated 1000 times per model. Empirical one-sided p-values were calculated using add-one smoothing as:

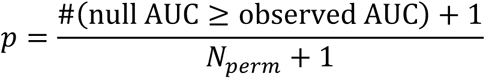

For computational tractability, the permutation analysis used reduced model settings and omitted RFE, because feature elimination on shuffled labels was not considered statistically meaningful.

### TreeSHAP interpretation

Random forest feature attributions were interpreted using TreeSHAP^41^. For each random-forest fold-fit, per-class and per-sample SHAP values were computed on the held-out test samples using SHAP. SHAP visualization was restricted to features selected by RFE in all fold-fits, ensuring that per-sample SHAP values were comparable across folds. SHAP values were concatenated across held-out folds before plotting.

### Statistical analysis

Statistical analyses were performed using Python (v3.8.8) and R (v4.2). Group comparisons were conducted using Mann-Whitney U tests or Kruskal–Wallis tests as appropriate. All statistical tests were two-sided, and p-values <0.05 after multiple-testing correction were considered statistically significant.

### Central adiposity measure analysis

All BMI association analyses were repeated for central-adiposity measures, with identical covariate adjustment for age, sex, and ethnicity. PERMANOVA (vegan adonis2, 9,999 permutations, Bray-Curtis) used the continuous measures (waist circumference, WHR, WHtR), each tested marginally and adjusted for the covariates (**Supplementary Table 1**). Differential abundance and supervised classification used the binary at-risk categories defined above (reference=normal), to parallel the categorical BMI analyses. Differential abundance was assessed with the same three tools as for BMI (MaAsLin2, ANCOM-BC2, DESeq2; **Supplementary Table 1**), and classification used the same pipeline as the taxonomic BMI classifier (species centred-log-ratio features, recursive feature elimination, 5×5 repeated stratified cross-validation, logistic-regression/linear-SVM/random-forest; **Supplementary Table 1**).

## Data availability

Metagenomic sequencing data has been deposited in the European Nucleotide Archive under accession number PRJEB115441. Source code and processed data required to reproduce figures and analyses are available under an MIT license at https://github.com/CSB5/helios_microbiome_obesity.

## Supplementary Tables

**Supplementary Table 1 |** Gut-microbiome associations with adiposity and host variables across three analytical frameworks. Results are provided as separate sheets: **PERMANOVA** variance explained in Bray-Curtis composition by BMI, waist circumference, WHR, WHtR and host factors (age, sex, ethnicity), marginally and covariate-adjusted; **cross-method differential-abundance** hit counts and overlap (MaAsLin2, ANCOM-BC2, DESeq2) by adiposity metric; and **supervised classification** performance by metric. All adiposity measures show near-null associations across frameworks. Full column definitions and per-analysis notes are given within each sheet.

**Supplementary Table 2 |** Differential abundance remains null within demographic subgroups. Species-level differential abundance (Obese vs Normal) was tested separately within ethnicity (Chinese) and sex (female, male) subgroups using three tools, MaAsLin2, ANCOM-BC2 and DESeq2, each adjusted for the remaining demographic covariates, with age modelled as a continuous term. For each subgroup the table reports the number of species reaching q<0.05 per tool, the number significant in both MaAsLin2 and ANCOM-BC2, the Fisher’s exact P-value for that cross-method overlap, and the identity of any overlapping species. In every stratum the conservative method (MaAsLin2) recovered at most one species, and no taxon was significant across two or more tools beyond chance expectation. The single MaAsLin2 ∩ ANCOM-BC2 overlap, in the Chinese subgroup (*Bifidobacterium adolescentis*), did not exceed chance (P = 0.18).

