## Supplementary Figures for "Population-scale analysis reveals limited and non-generalizable associations between the gut microbiome and obesity in Asian adults"

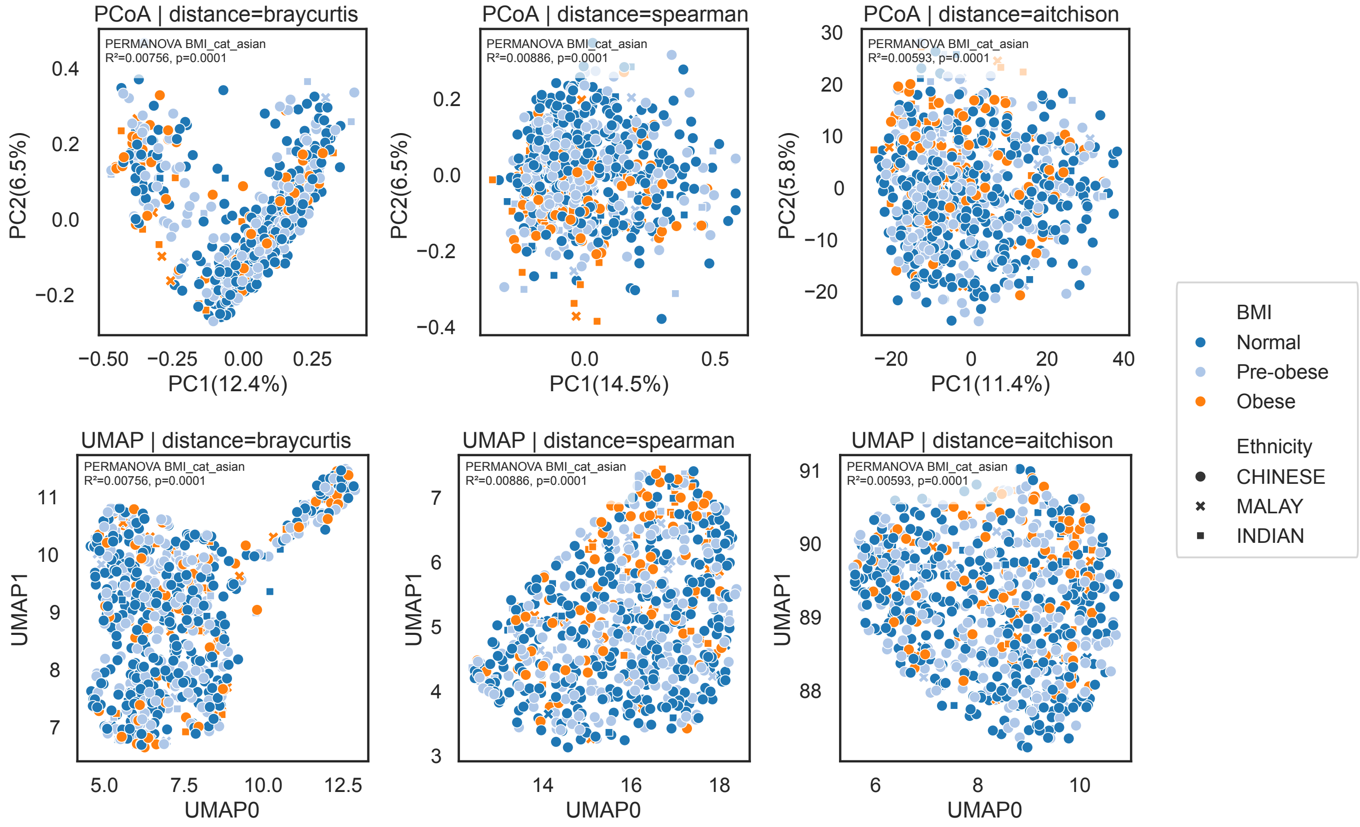


**Supplementary Figure 1 | Ordination of gut microbiome composition across alternative distance metrics shows weak separation by BMI category.** Principal coordinates analysis (PCoA, top row) and Uniform Manifold Approximation and Projection (UMAP, bottom row) of species-level gut microbiome profiles using Bray-Curtis, Spearman, and Aitchison distances. Samples are colored by Asian-specific BMI category (Normal, Pre-obese, Obese) and marked with shapes corresponding to self-reported ethnicity (Chinese, Malay, Indian). Across all distance metrics and ordination methods, samples show broad overlap with no distinct clustering by BMI category or ethnicity. PERMANOVA results for BMI are shown within each panel and indicate that BMI explains only a very small fraction of community variation across distance metrics (<1%).

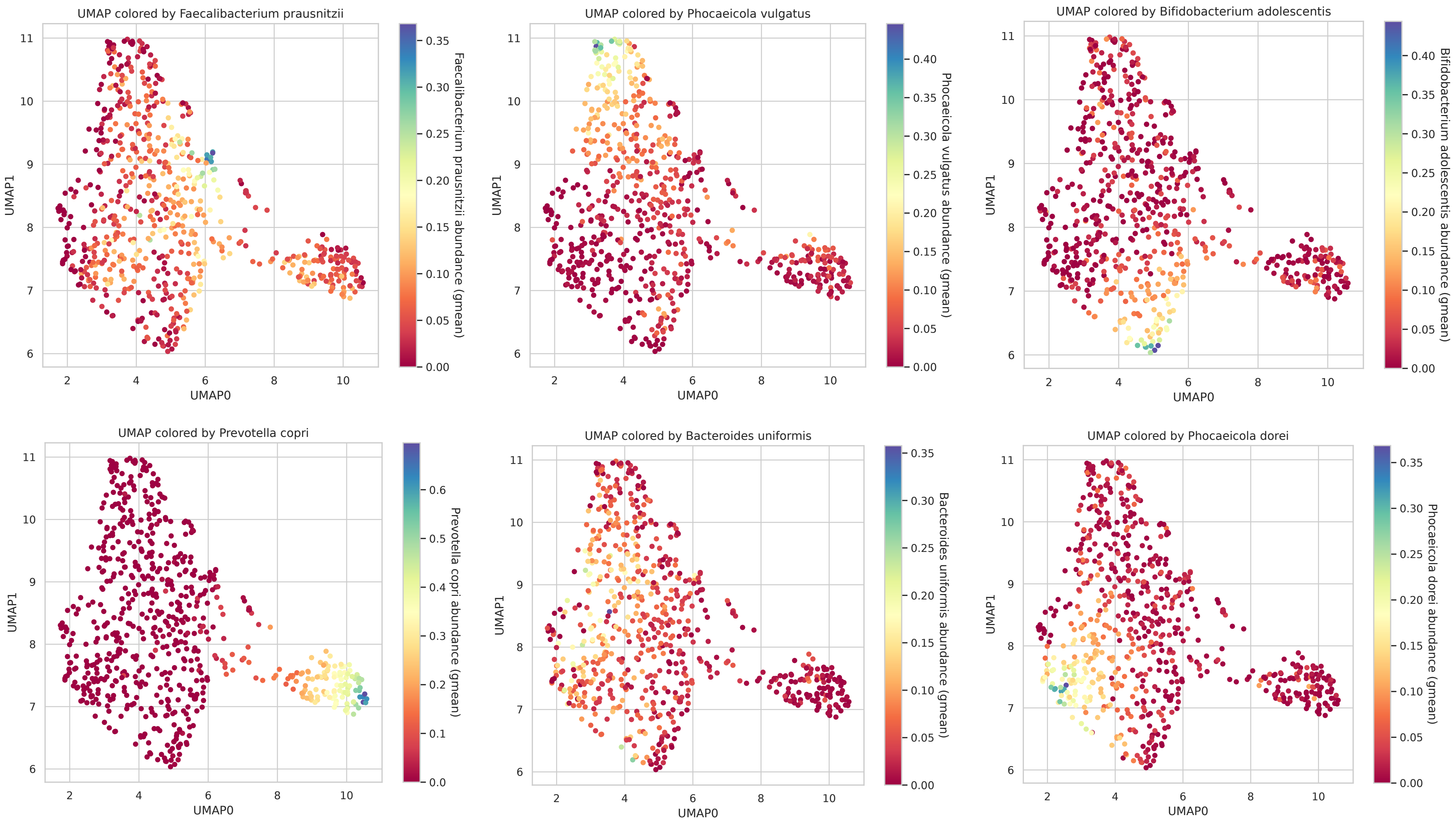


**Supplementary Figure 2 | Uniform Manifold Approximation and Projection (UMAP) of species-level gut microbiome profiles using Bray–Curtis distance.** Each plot is colored by the abundance of a key species that is enriched in the six different clusters identified.

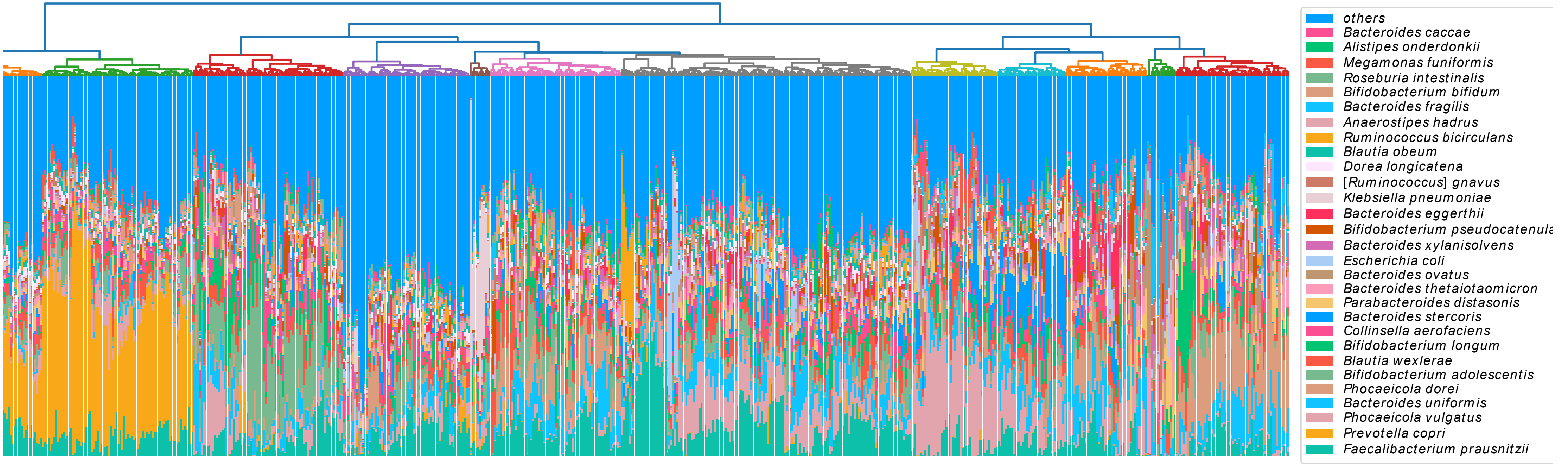


**lementary Figure 3 | Hierarchical clustering of species-level gut microbiome profiles.** Stacked bar plots show the relative abundances of the most abundant species across les, with remaining taxa grouped as “others.” Samples are ordered according to hierarchical clustering based on species-level community composition, and the dendrogram ırity relationships among samples. The figure highlights broad gradients in the relative abundance of prevalent gut commensals across the cohort.

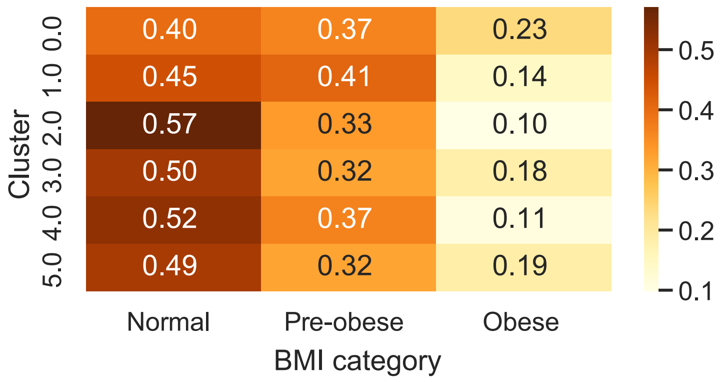


**Supplementary Figure 4 | Distribution of BMI categories across spectral clusters.** Heatmap showing the proportion of samples in each BMI category (Normal, Pre-obese, Obese) within each spectral cluster. Values in each row sum to 1 and represent within-cluster proportions. Although some clusters show modest differences in category composition, the overall distribution of BMI categories across clusters was not significantly different ($\chi^2$ = 16.52, P = 0.086), indicating that there was no strong enrichment for obesity within any single cluster.

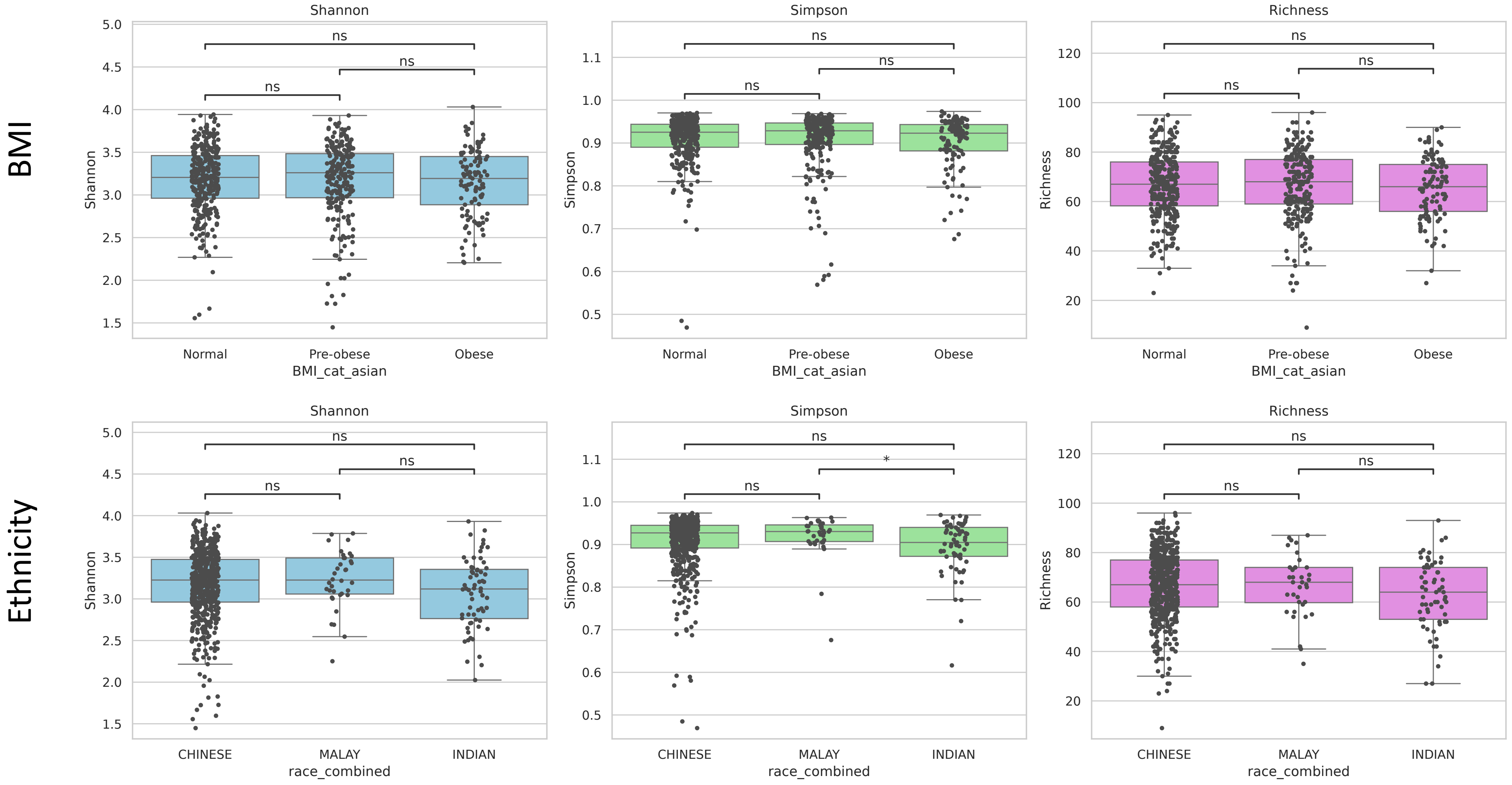


**Supplementary Figure 5 | Alpha diversity across BMI categories and ethnic groups.** Boxplots showing Shannon diversity, Simpson diversity, and richness stratified by Asian-specific BMI category (Normal, Pre-obese, Obese; top row) and self-reported ethnicity (Chinese, Malay, Indian; bottom row). Points represent individual samples. Pairwise comparisons are annotated above each panel, with significance indicated as shown (ns, not significant; *, adjusted *P*<0.05). Overall, alpha diversity metrics were broadly similar across BMI categories, whereas only limited differences were observed across ethnic groups.

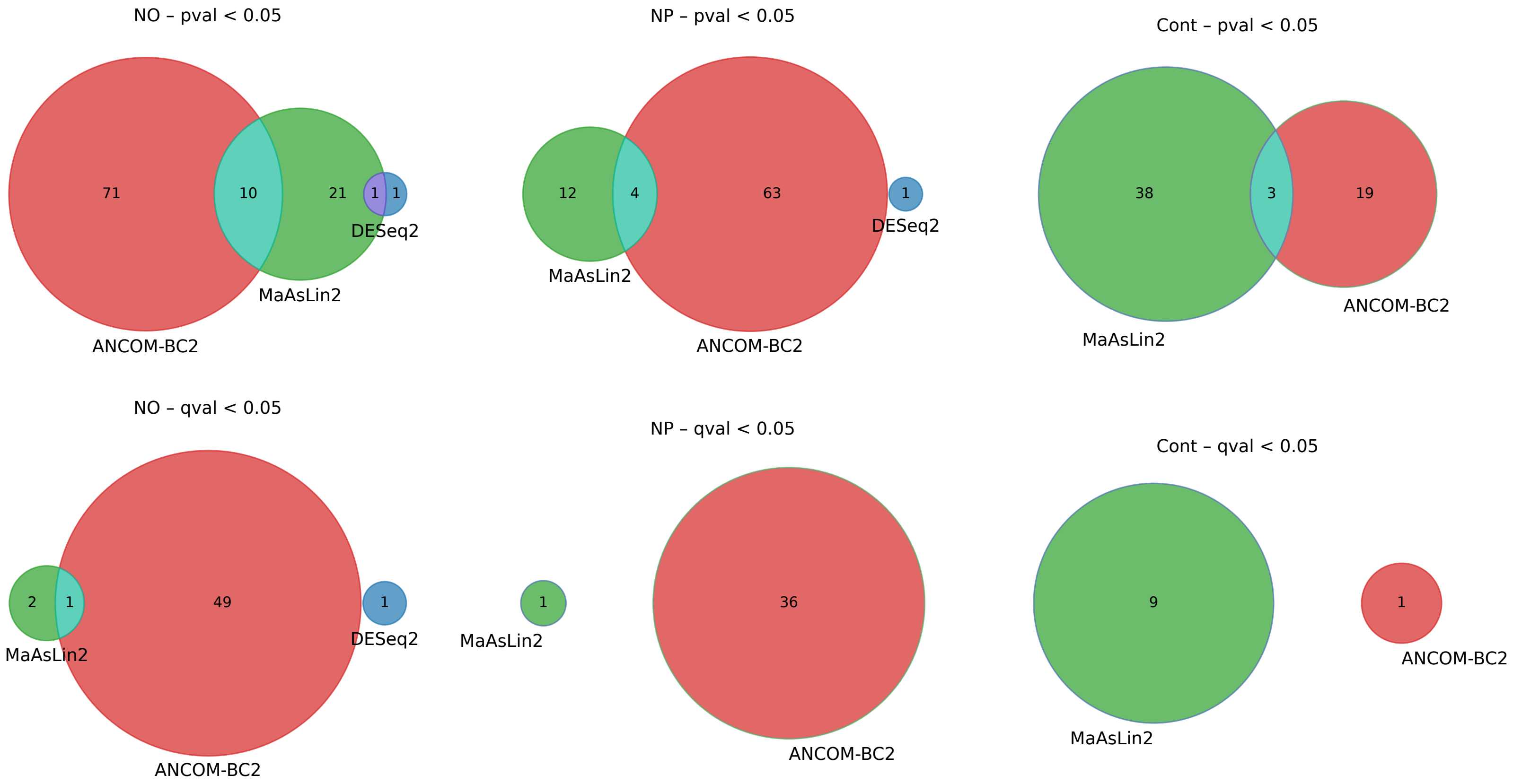


**Supplementary Figure 6 | Cross-method overlap of taxonomic differential abundance signals across BMI contrasts.** Venn diagrams showing the overlap of taxa identified as differentially abundant by ANCOM-BC2, MaAsLin2, and DESeq2 across three BMI comparisons: Normal vs. Obese (NO), Normal vs. Pre-obese (NP), and BMI as a continuous trait (Cont). The top row shows taxa passing a nominal threshold (P<0.05), whereas the bottom row shows taxa remaining significant after multiple-testing correction (q<0.05). Across all contrasts, overlap between methods was limited, with most significant taxa identified by only one framework. Cross-method agreement was greatest for the normal vs. obese contrast, but remained sparse after multiple-testing correction.

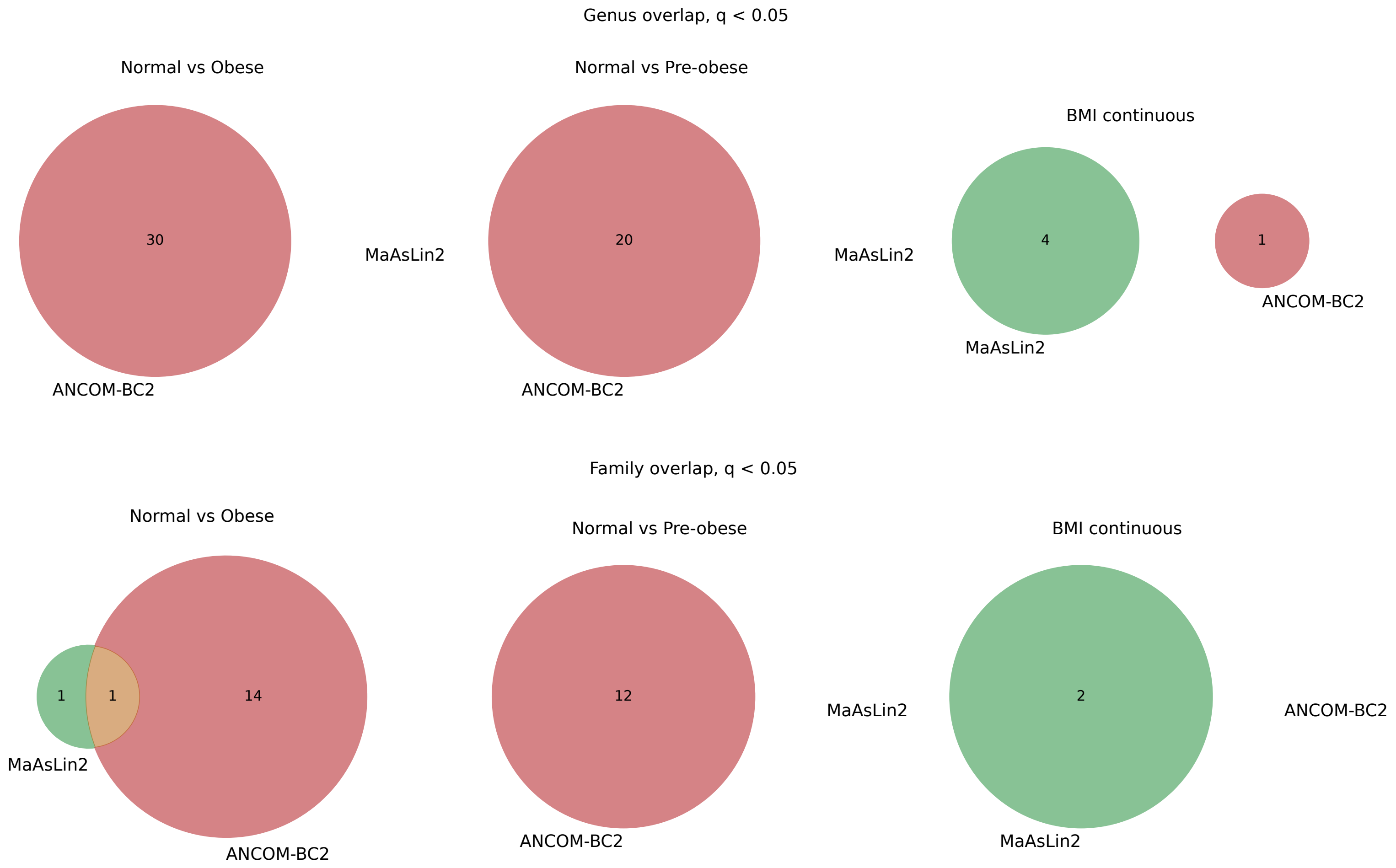


**Supplementary Figure 7 | Limited cross-method concordance of differential abundance results at the genus and family levels.** Venn diagrams showing overlap between ANCOM-BC2, MaAsLin2 and DESeq2 (which had no hits) for taxa identified as differentially abundant at the genus level (top row) and family level (bottom row) across three phenotype definitions: Normal vs. Obese, Normal vs Pre-obese, and BMI as a continuous trait. Only taxa passing multiple-testing correction (q<0.05) are shown. Across both taxonomic levels, overlap between methods remained limited, with the strongest signal observed for the Normal vs. Obese contrast and little or no concordance for the other comparisons.

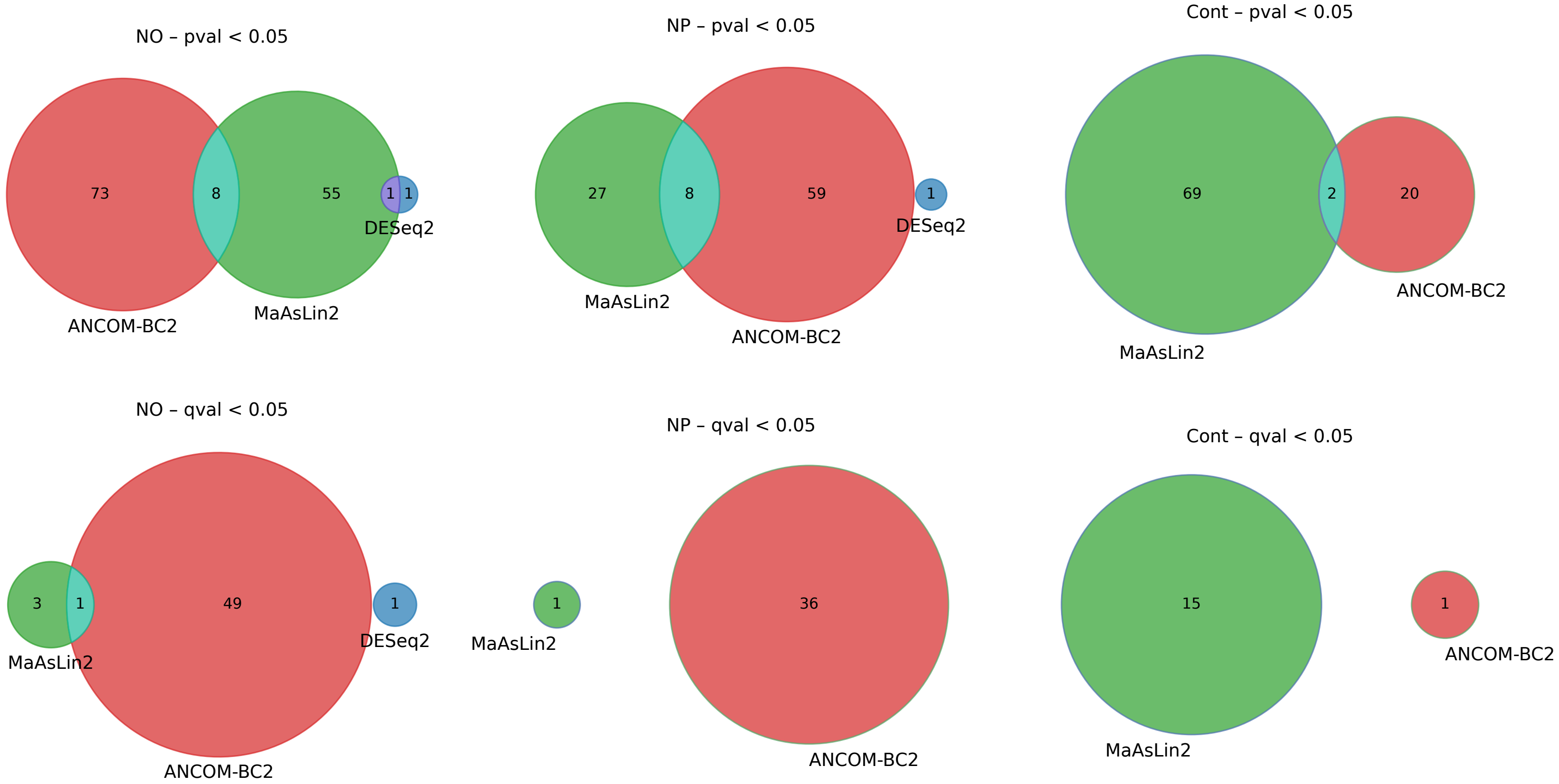


**Supplementary Figure 8 | Relaxed abundance filtering of 0.01% increases taxonomic hits without improving cross-method concordance.** Venn diagrams showing overlap among taxa identified by ANCOM-BC2, MaAsLin2, and DESeq2 across the three BMI contrasts after relaxing the relative-abundance filter. Top row shows nominal P < 0.05; bottom row shows FDR-adjusted q < 0.05. Although moderately more taxa were detected at nominal significance, overlap across methods remained limited, and little additional concordance was observed after multiple-testing correction. This suggests that relaxing the filter mainly admits extra low-abundance, method-specific associations rather than strengthening robust shared signals.

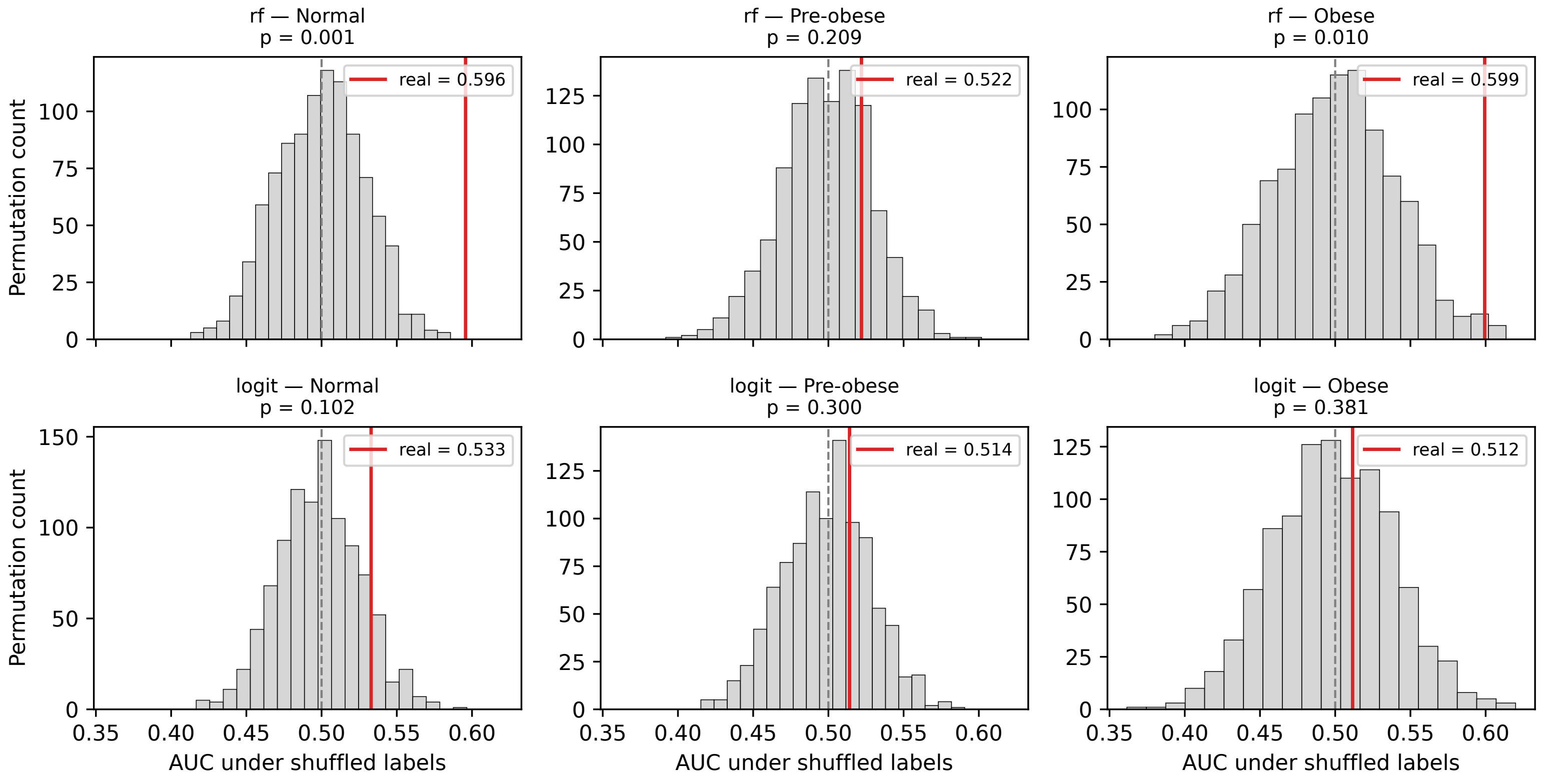


**Supplementary Figure 9 | Permutation-label null distributions and empirical significance of per-class AUC-ROC.** For each of two reference classifiers (random forest, top row; l2-penalised logistic regression, bottom row), BMI labels were shuffled across the cohort and the classifier was refit under the 5-fold stratified cross-validation as the main analyses, yielding one null AUC per class per permutation; N = 1000 permutations per (model, class) combination. Each panel shows the null AUC histogram (grey, 1000 values), the observed real AUC for that (model, class) pair (red vertical line), and the chance line at AUC = 0.5 (dashed grey). Empirical p-values were computed as the fraction of permutations with null AUC ≥ real AUC, with add-one smoothing. To isolate the contribution of the input matrix from feature selection, the reference classifiers used the full CLR matrix without RFE. The random forest reached statistical significance against the null for the Normal class (real AUC = 0.596, p = 0.001) and the Obese class (real AUC = 0.599, p = 0.010), but not for Pre-obese (real AUC = 0.522, p = 0.21). The logistic regression model showed no significant signal at any class (Normal p = 0.10, Pre-obese p = 0.30, Obese p = 0.38).

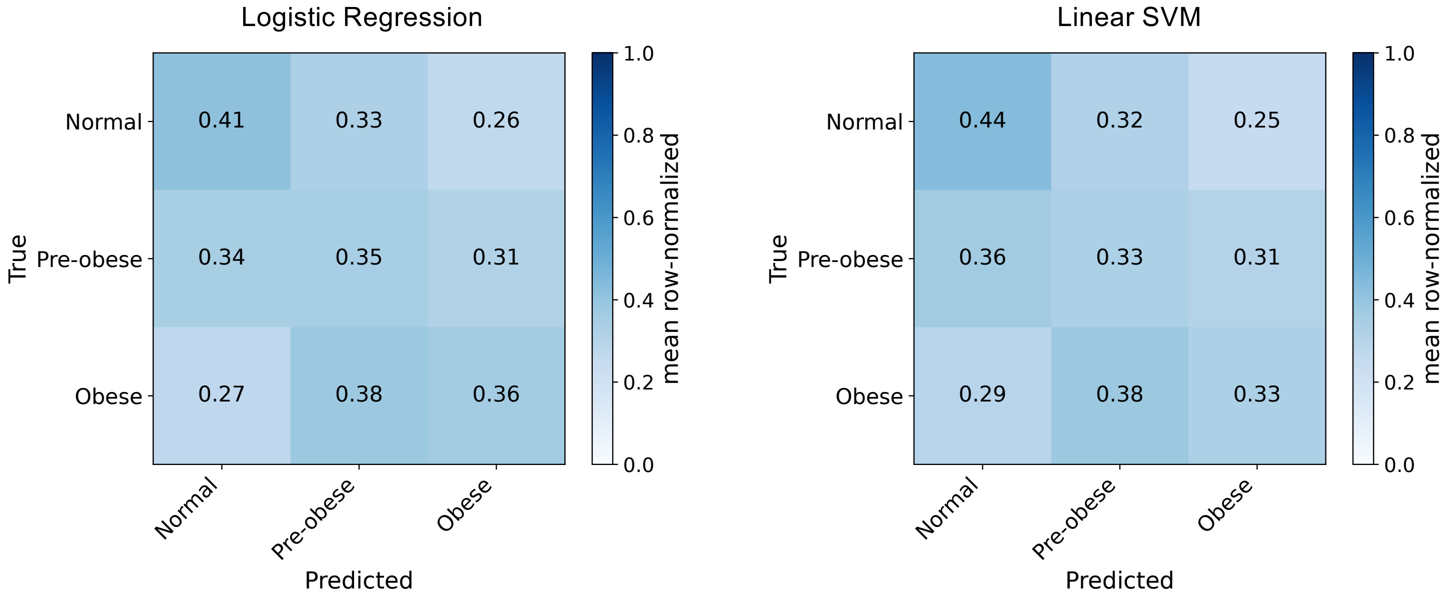


**Supplementary Figure 10 | Mean normalized confusion matrices for the two linear classifiers, complementing the random forest panel in Fig. 3C.** Logistic regression (**left**) and linear-kernel support vector machine (**right**). Cells are row-normalized (rows = true class, columns = predicted class) and averaged across 25 fold-fits (5-fold stratified cross-validation × 5 repeats). Both linear classifiers distributed their predictions diffusely across all three BMI classes rather than concentrating on the correct diagonal. Mean diagonal accuracies, logit: 0.43 (Normal) / 0.35 (Pre-obese) / 0.37 (Obese); linear SVM: 0.44 / 0.33 / 0.33, are near the chance baseline of 1/3 for a 3-class problem. Off-diagonal mass is approximately symmetric: errors did not preferentially fall on adjacent BMI classes (Normal misclassified as Pre-obese versus Obese: 0.33 vs. 0.24 for logit, 0.32 vs. 0.25 for SVM), ruling out a "noisy-ordinal" interpretation of the near-random performance. The diffuse pattern stands in contrast to the random forest's majority-class collapse (Fig. 3C), in which RF classifies 75–86% of all samples as Normal despite balanced class weighting; the three models therefore fail in qualitatively different ways while achieving indistinguishable mean diagonal accuracy.

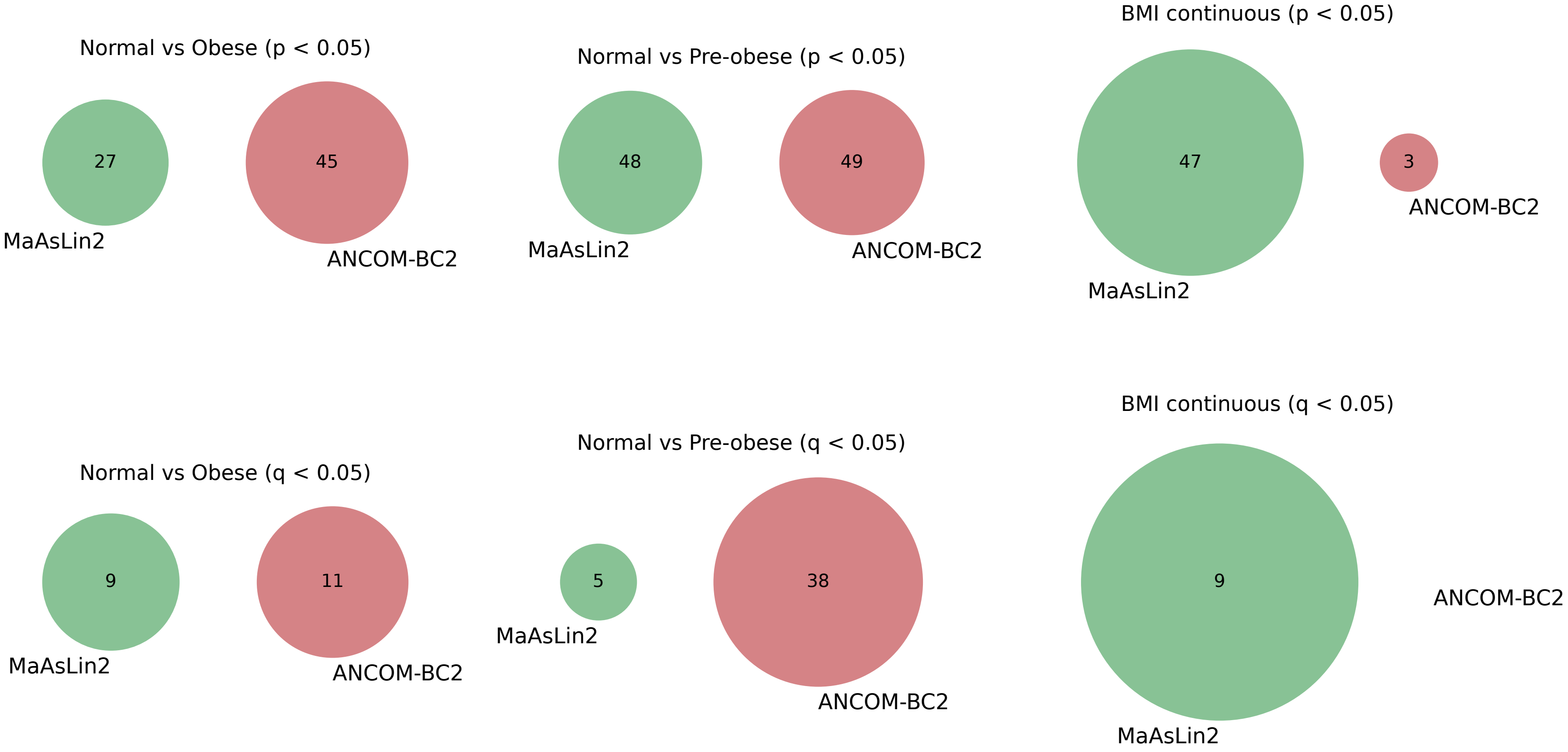


**Supplementary Figure 11 |Concordance of MetaCyc pathway associations between MaAsLin2 and ANCOM-BC2 across BMI contrasts.** Venn diagrams show the overlap (or lack thereof) of differentially abundant MetaCyc pathways identified by MaAsLin2 (green) and ANCOM-BC2 (pink) across three BMI contrasts (Normal vs. Obese, Normal vs. Pre-obese, BMI continuous; columns) and two significance thresholds (nominal $p < 0.05$, top row; FDR-adjusted $q < 0.05$, bottom row). Circle area is proportional to the number of significant pathways. DESeq2 is not shown as it does not natively account for the compositional structure of metagenomic pathway abundance data and was therefore not applied in this analysis; MaAsLin2 and ANCOM-BC2 were selected as the primary methods given their established suitability for compositional microbiome data. At the nominal threshold, both methods detected large pathway sets (up to 49 pathways), yet concordance was negligible across all contrasts. After FDR correction, the total number of significant hits contracted markedly - particularly for ANCOM-BC2 under the BMI-continuous contrast (3 pathways at $p < 0.05$; 0 surviving $q < 0.05$) - and inter-method overlap remained absent. All analyses were adjusted for age, sex, and race.

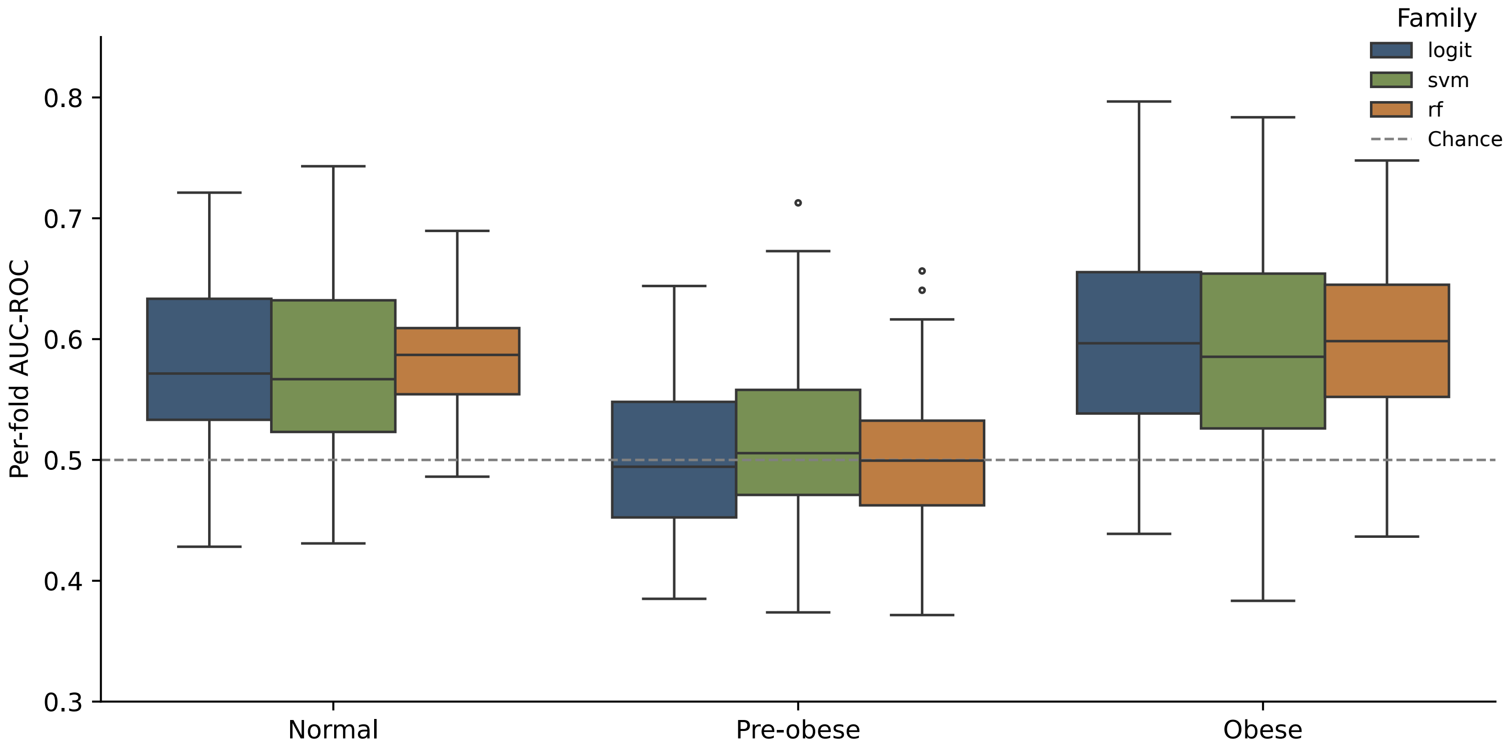


**Supplementary Figure 12 | Per-class AUC-ROC for BMI classification from MetaCyc pathways is near random across three classifier families.** Per-fold AUC-ROC (y-axis) for classification of each BMI category (Normal, Pre-obese, Obese) using three classifier families on the unstratified MetaCyc pathway abundance matrix: elastic-net logistic regression (logit; blue), linear-kernel support vector machine (SVM; green), and random forest (RF; orange). Models were fitted under 5-fold × 10-repeat stratified cross-validation with recursive feature elimination to K = 50 features applied independently inside each outer training fold; each box summarises 25 fold-fits (5 folds × 5 repeats). The dashed grey line marks chance performance (AUC = 0.5). Across all three families the Pre-obese class is indistinguishable from chance (median AUC ≈ 0.50; whiskers spanning ≈ 0.37–0.71), while the Normal and Obese extremes reach only modestly above chance (median ≈ 0.58–0.60), with per-fold spread crossing the chance line in every cell.

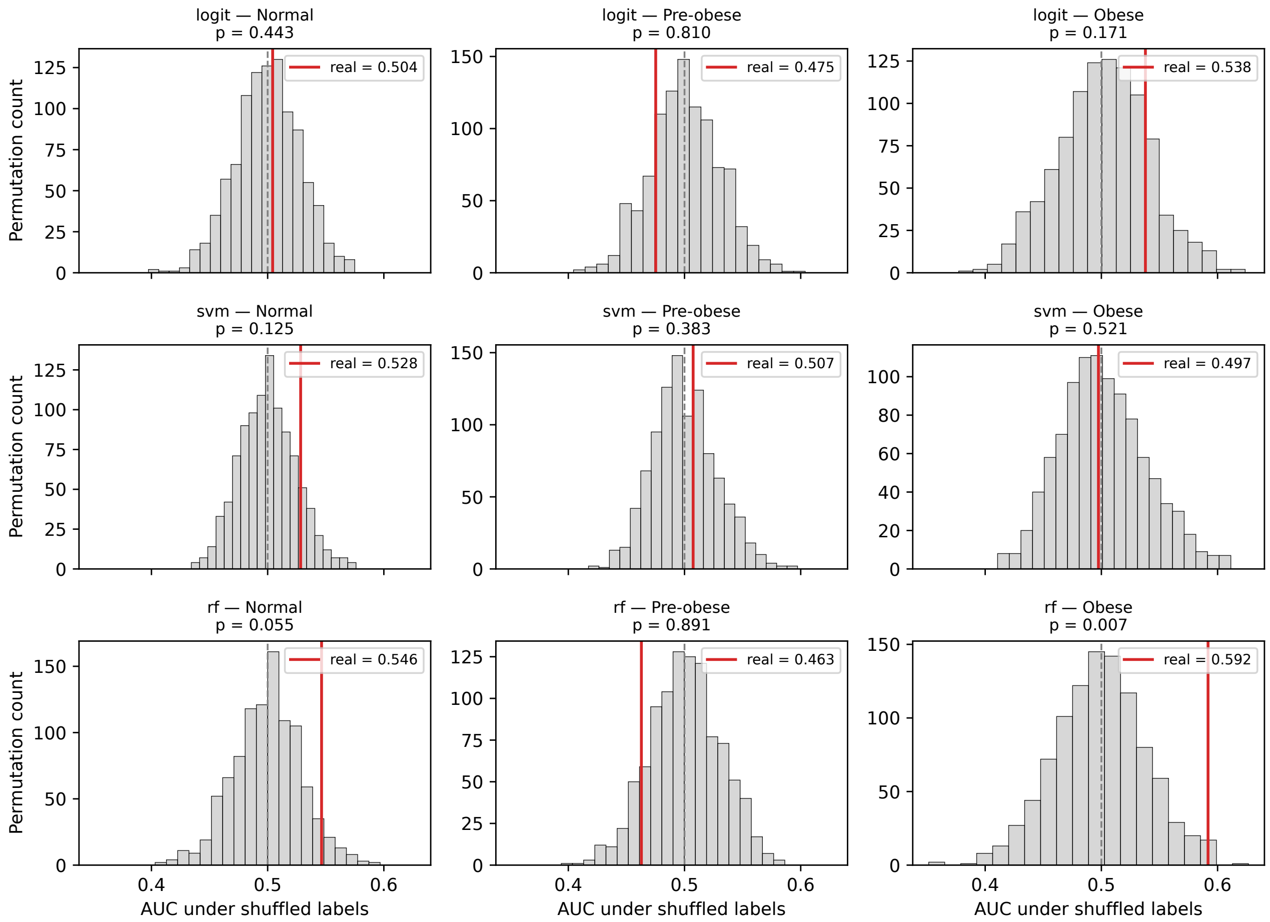


**Supplementary Figure 13 | Permutation null distributions for MetaCyc one-vs-rest classifiers.** Each panel shows the empirical null distribution of one-vs-rest AUC-ROC obtained from 200 label-permutation replicates (grey histograms) for three classifier families (rows: Elastic-Net Logistic Regression, Linear SVM, Random Forest) and three BMI classes (columns: Normal, Pre-obese, Obese). The dashed grey line marks the mean of the null distribution; the red vertical line indicates the observed AUC from the true labels. Permutation p-values (proportion of null AUCs ≥ observed AUC) are shown above each panel. All observed AUCs fall within or near the permutation null, with no comparison surviving a Bonferroni correction for nine tests ($\alpha = 0.05/9 \approx 0.006$); the nominally lowest p-value (RF – Obese, $p = 0.007$) does not meet this threshold. These results confirm that MetaCyc functional pathway profiles carry no statistically significant discriminative signal for BMI category in this cohort.

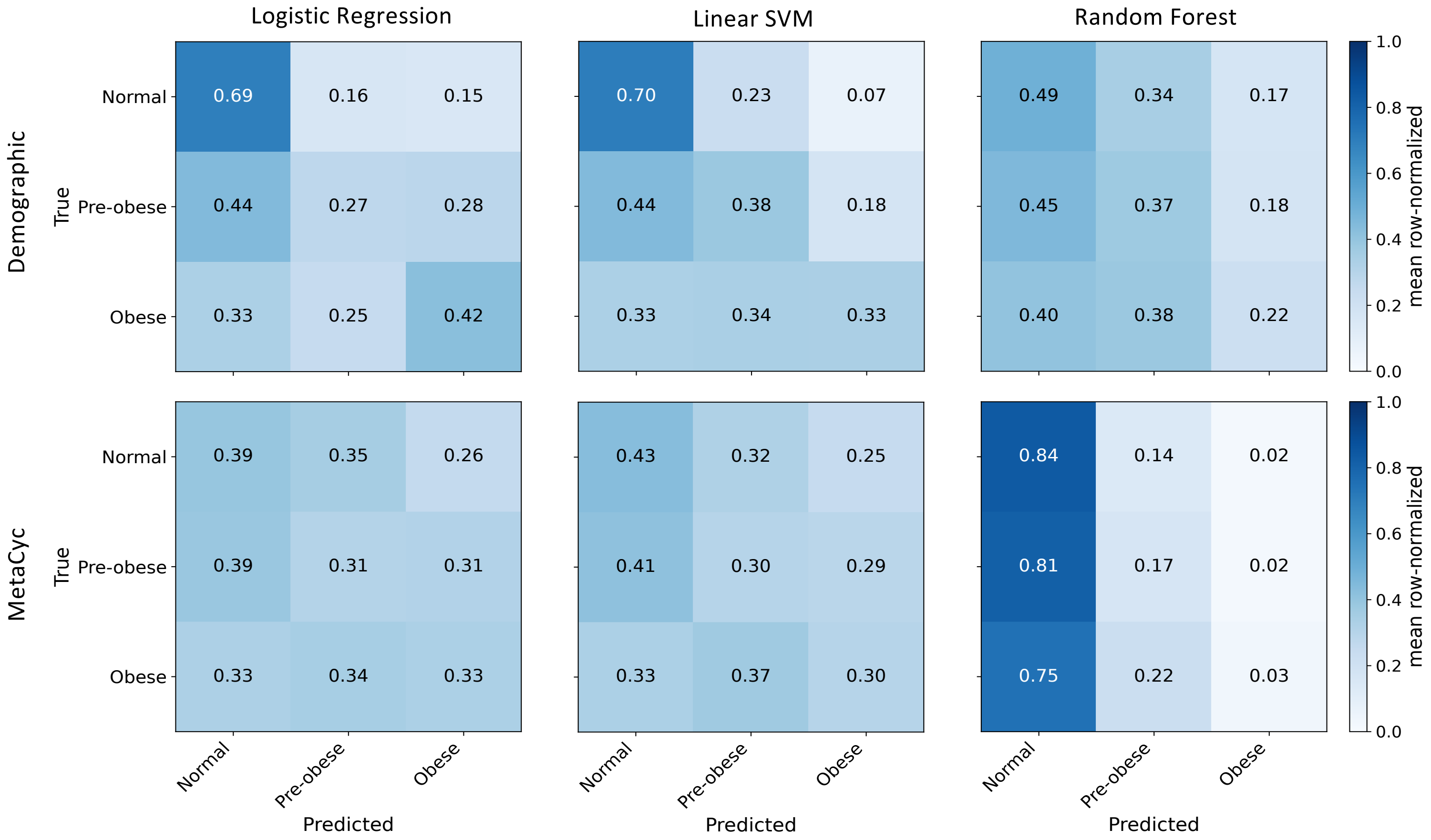


**Supplementary Figure 14 | Mean normalized confusion matrices for demographic and MetaCyc classifiers.** Row-normalized confusion matrices averaged across 25 cross-validation folds (5 repeats × 5 folds) for three-class BMI prediction (Normal, Pre-obese, Obese). Top row: classifiers trained on demographic features; bottom row: classifiers trained on MetaCyc functional pathway relative abundances. Columns show three classifier families: Elastic-Net Logistic Regression (logit), Random Forest (RF), and Linear SVM. Cell values indicate the mean proportion of true-class samples assigned to each predicted class; a perfect classifier would show 1.0 on the diagonal. Demographic classifiers achieve modest diagonal enrichment, particularly for the Normal class (logit: 0.69, SVM: 0.70), whereas MetaCyc classifiers perform near chance level (~0.33 per cell), with RF collapsing almost entirely to a Normal-prediction bias (diagonal values 0.84 / 0.17 / 0.03 for true Normal / Pre-obese / Obese, respectively). The Pre-obese class is the most difficult to recover across all feature sets and classifiers, with substantial misclassification as Normal. Together, these matrices confirm that MetaCyc functional profiles add no discriminative information beyond random assignment for BMI category classification.

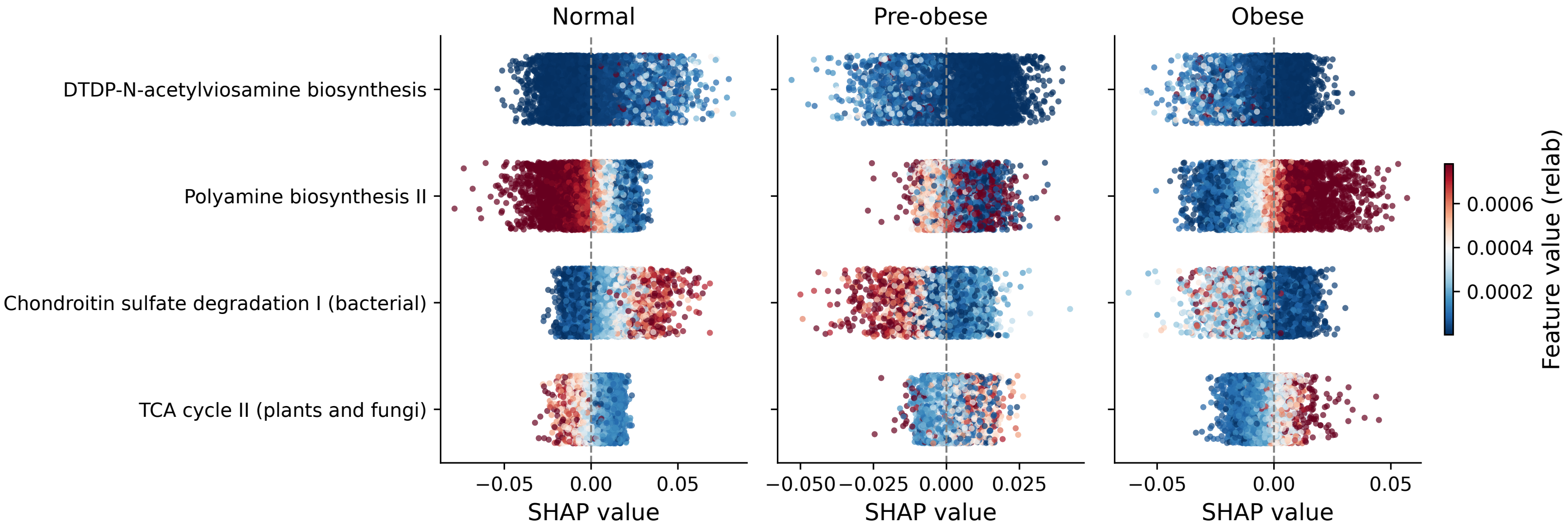


**Supplementary Figure 15 | TreeSHAP beeswarm plots for the Random Forest MetaCyc classifier.** Each panel shows per-sample SHAP values for one-vs-rest classification of a BMI class (Normal, Pre-obese, Obese) using MetaCyc functional pathway relative abundances. Only the top features passing a 100% cross-validation stability filter are shown (top 4 displayed here for clarity). Each dot represents one sample from one cross-validation fold; dot colour encodes the feature's relative abundance (blue = low, red = high; scale on right). Positive SHAP values indicate a contribution toward predicting membership in that class; negative values push the prediction away. The SHAP magnitudes are uniformly small (|SHAP| < 0.075) and no pathway shows a consistent directional effect across all three classes, confirming the absence of a robust functional signature of BMI category. Polyamine biosynthesis II exhibits the clearest gradient (higher abundance associated with reduced Normal and Pre-obese probability and modestly increased Obese probability), but its effect size is negligible and inconsistent with a discriminating biomarker.

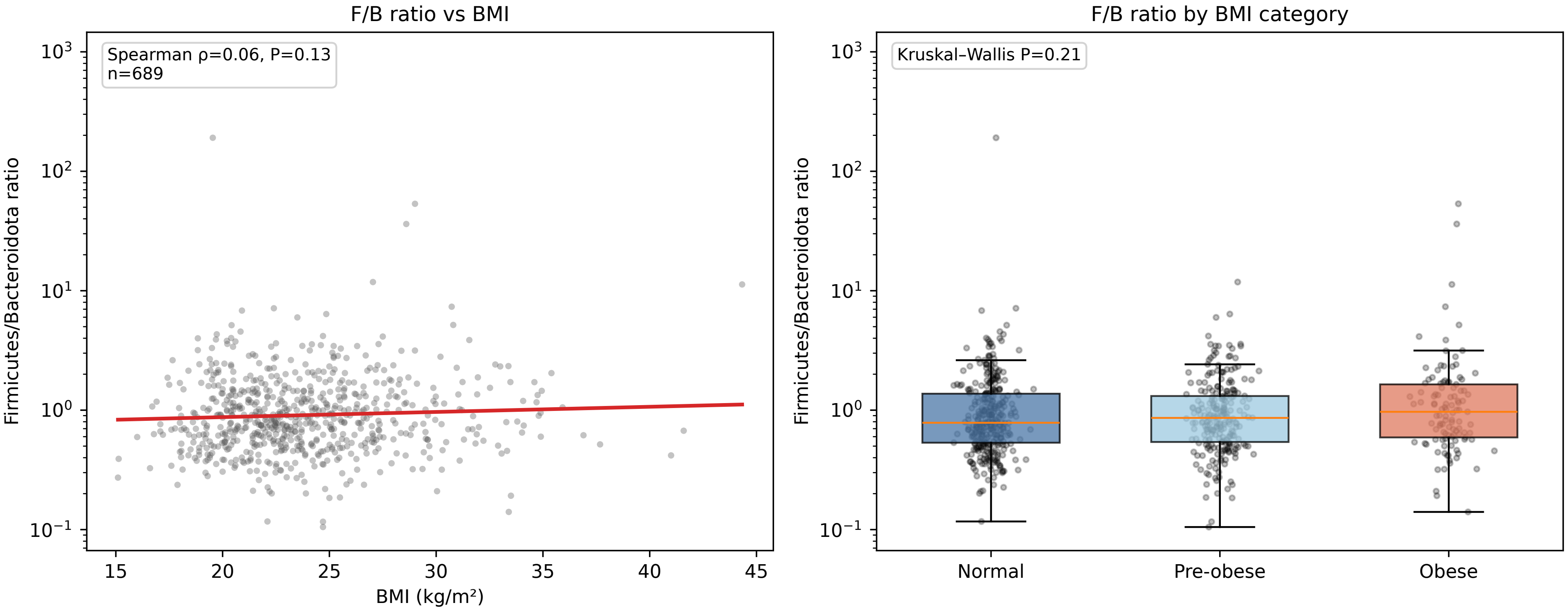


**Supplementary Figure 16 |** Firmicutes-to-Bacteroidota (F/B) ratio versus BMI. F/B ratio (phylum-level relative abundances, $\log_{10}$ scale) shown against continuous BMI (left; red line, linear fit; Spearman ρ = 0.06, P = 0.13; n = 689) and across BMI categories (right; box, median and interquartile range; whiskers, 1.5× IQR; Points represent individual participants.) Neither comparison is significant.
